# Risk factors for long covid in previously hospitalised children using the ISARIC Global follow-up protocol: A prospective cohort study

**DOI:** 10.1101/2021.04.26.21256110

**Authors:** Ismail M Osmanov, Ekaterina Spiridonova, Polina Bobkova, Aysylu Gamirova, Anastasia Shikhaleva, Margarita Andreeva, Oleg Blyuss, Yasmin El-Taravi, Audrey DunnGalvin, Pasquale Comberiati, Diego G Peroni, Christian Apfelbacher, Jon Genuneit, Lyudmila Mazankova, Alexandra Miroshina, Evgeniya Chistyakova, Elmira Samitova, Svetlana Borzakova, Elena Bondarenko, Anatoliy A Korsunskiy, Irina Konova, Sarah Wulf Hanson, Gail Carson, Louise Sigfrid, Janet T Scott, Matthew Greenhawt, Elizabeth A Whittaker, Elena Garralda, Olivia Swann, Danilo Buonsenso, Dasha E Nicholls, Frances Simpson, Christina Jones, Malcolm G Semple, John O Warner, Theo Vos, Piero Olliaro, Daniel Munblit, Sechenov StopCOVID Research Team

## Abstract

**Background:** The long-term sequelae of coronavirus disease 2019 (Covid-19) in children remain poorly characterised. This study aimed to assess long-term outcomes in children previously hospitalised with Covid-19 and associated risk factors.

**Methods:** This is a prospective cohort study of children (≤18 years old) admitted with confirmed Covid-19 to Z.A. Bashlyaeva Children’s Municipal Clinical Hospital in Moscow, Russia. Children admitted to the hospital during the first wave of the pandemic, between April 2, 2020 and August 26, 2020, were included. Telephone interview using the International Severe Acute Respiratory and emerging Infection Consortium (ISARIC) Covid-19 Health and Wellbeing paediatric follow up survey. Persistent symptoms (>5 months) were further categorised by system(s) involved.

**Findings:** Overall, 518 of 853 (61%) of eligible children were available for the follow-up assessment and included in the study. Median age was 10.4 years (IQR, 3–15.2) and 270 (52.1%) were girls; median follow-up since hospital discharge was 256 (223-271) days. At the time of the follow-up interview 126 (24.3%) participants reported persistent symptoms among which fatigue (53, 10.7%), sleep disturbance (36, 6.9%,) and sensory problems (29, 5.6%) were the most common. Multiple symptoms were experienced by 44 (8.4%) participants. Risk factors for persistent symptoms were: age “6-11 years” (odds ratio 2.74 (95% confidence interval 1.37 to 5.75) and “12-18 years” (2.68, 1.41 to 5.4), and a history of allergic diseases (1.67, 1.04 to 2.67).

**Interpretation:** A quarter of children experienced persistent symptoms months after hospitalization with acute covid-19 infection, with almost one in ten experiencing multi-system involvement. Older age and allergic diseases were associated with higher risk of persistent symptoms at follow-up. Our findings highlight the need for replication and further investigation of potential mechanisms as well as clinical support to improve long term outcomes in children.

**Funding:** None.

Research in context

Evidence before this study
Evidence suggests that Covid-19 may result in short- and long-term consequences to health. Studies in children and adolescents are limited and available evidence is scarce. We searched Embase for publications from inception to April, 25, 2021, using the following phrases or combinations of phrases “post-covid condition” or “post-covid syndrome” or “covid sequalae” or “post-acute covid” or “long covid” or “long hauler” with “pediatric*” or “paediatric*” or “child*” or “infant*” or “newborn*” or “toddler*” or “neonate*” or “neonatal” or “adolescent*” or “teen*”. We found small case series and small cohort studies looking at Covid-19 consequences in children. No large cohort studies of previously hospitalised children, assessing symptom duration, categorisation or attempting multivariable analyses to identify independent risk factors for long Covid development were identified.

Added value of this study
To our knowledge, this is the largest cohort study with the longest follow-up since hospital discharge of previously hospitalised children. We found that even months after discharge from the hospital, approximately a quarter of children experience persistent symptoms with one in ten having multi-system involvement. Older age and allergic diseases are associated with Covid-19 consequences. Parents of some children report emotional and behavioural changes in their children after Covid-19.

Implications of all the available evidence
Our findings highlight the need for continued global research of Covid-19 consequences in the paediatric population. Older children admitted to the hospital should be carefully monitored upon discharge. Large, controlled studies aiming to identify risk groups and potential intervention strategies are required to fill knowledge gaps.

## INTRODUCTION

The Covid-19 pandemic has, at the time of writing, affected over 135 million people worldwide ^1^, with adverse impacts on physical and psychological health and over 3 million deaths. While our knowledge of the acute phase of the disease has increased over time, evidence on the longer-term health consequences of covid-19 is still limited. Emerging data suggest that a substantial proportion of people experience ongoing symptoms including fatigue and muscle weakness, breathlessness, and neurological problems more than 6 months after the acute phase ^2,3^. This phenomenon is commonly referred to as ‘long Covid’, a term defined by patient groups, and also known as post-Covid syndrome, the post-Covid-19 condition ^4^ or ‘Covid long-haulers ^5,6^. Recent population data from the UK reported that the highest prevalence of long Covid after 12 weeks was among those aged 25 to 34 years (18.2%) and lowest in the 2 to 11 years age bracket (7.4%)^7^.

Children and adolescents are at lower risk of severe Covid-19 illness compared to adults ^8^ and may present with a wide range of clinical features and symptoms at the time of hospital admission ^9-11^. Data suggest that among children hospitalised with Covid-19, up to a third of patients require intensive care (ICU) ^12^ with premature infants at higher risk^10^. Among children admitted to ICU, some experience single or multi-organ failure and many require respiratory support. A small number of patients develop a severe condition called multisystem inflammatory syndrome in children (MIS-C), which usually appears a few weeks after the acute infection ^13^. Evidence on post-acute covid condition and long term outcomes in children is still limited to small studies with more than half having at least one persisting symptom 4 months after covid-19 infection ^14^. However, a recent publication from Australia suggested that only 8% of children aged 0-19 years (median 3 years) had ongoing symptoms 3-6 months after predominantly mild covid-19 infection. The limitation of the study as acknowledged by the authors was the low age range. This mandates the inclusion larger numbers particularly of older children in future studies ^15^.

There is a need to assess the long-term consequences of Covid-19 in paediatric populations ^16^, to inform clinicians, researchers and public health experts and address the impacts of this condition on the quality of life (QoL) of those affected and their families and to inform discussions on vaccination of children. This cohort study aimed to investigate the incidence of and risk factors for long-term Covid-19 outcomes in children post-hospital discharge. We used the standardised follow-up data collection protocol developed by the International Severe Acute Respiratory and Emerging Infection Consortium (ISARIC) Global Paediatric Covid-19 follow up working group ^17^.

## METHODS

### Study design, setting and participants

This is a prospective cohort study of children (≤18 years old) admitted with suspected or confirmed Covid-19 to Z.A. Bashlyaeva Children’s Municipal Clinical Hospital in Moscow, Russia. This large tertiary university hospital can accommodate up to 980 children at a time and served as the primary Covid-19 hospital for children residing in Moscow city. Children admitted to the hospital during the first wave of the pandemic, between April 2, 2020 and August 26, 2020, with reverse transcriptase polymerase chain reaction (RT-PCR) confirmed SARS-CoV-2 infection were included. The parents of these children were contacted between January 31, 2021 and February 27, 2021 to complete a follow up survey for this study.

The acute-phase data were extracted from electronic medical records (EMR) and the Local Health Information System (HIS) at the host institution. The acute-phase dataset included demographics, symptoms, co-morbidities, chest computer tomography (CT), supportive care, and clinical outcomes at discharge. This study was approved by the Moscow City Independent Ethics Committee (abbreviate 1, protocol number 74). Parental consent was sought during hospital admission and consent for the follow-up interview was sought via verbal confirmation during telephone interview.

Information about the current condition and persisting symptoms was collected by trained medical students via telephone by interviewing the parent/carer of the child, using the version 1 of the ISARIC COVID-19 Health and Wellbeing Follow Up Survey for Children, to assess patients’ physical and psychosocial wellbeing and behaviour, with local adaptations, translated into Russian. The protocol was registered at The Open Science Framework ^18^. The follow-up survey documented data on demographics, parental perception of changes in their child’s emotional and behavioural status, previous vaccination history, hospital stay and readmissions, mortality (after the initial index event), history of newly developed symptoms between discharge and the follow-up assessment, including symptom onset and duration, and overall health condition compared to prior to the child’s Covid-19 onset (**Supplementary file**). To assess the prevalence of symptoms over time parents were asked the following: (a) *Within the last seven days, has your child had any of these symptoms, which were NOT present prior to their Covid-19 illness? (If yes, please indicate below and the duration of the symptom/s) and (b) Please report any symptoms that have been bothering your child since discharge that are not present today. Please specify the time of onset and duration of these symptoms*.

Interviews were undertaken by a team of medical students with experience gained in previous Covid-19 research ^3,19^ who underwent standardised training in telephone assessment, REDCap data entry and data security. Assessments were conducted via interviews with the parents/carers. Non-responders were contacted by telephone three times before considering them lost to follow up.

### Data management

REDCap electronic data capture tools (Vanderbilt University, Nashville, TN, USA) hosted at Sechenov University and Microsoft Excel (Microsoft Corp, Redmond, WA, USA) were used for data collection, storage and management ^20,21^. The baseline characteristics, including demographics, symptoms on admission and comorbidities were extracted from EMRs and entered into REDCap.

### Exposure and outcome variables

For the purposes of this study, we defined “persistent symptoms” as symptoms present at the time of the follow-up interview and lasting for over 5 months. These were subcategorised into respiratory, neurological, sensory, sleep, gastrointestinal, dermatological, cardiovascular, fatigue and musculoskeletal (Table S1) informed by previously published literature ^22,23^ and international expert group discussions.

Allergic diseases were defined as a presence of any of the following: asthma, allergic rhinitis, eczema or food allergy. Health status before Covid-19 and at the time of the interview was assessed using a 0 to 100 wellness scale ^24^, where 0 was the worst possible health and 100 the best possible health. Participants age categories were based on Eunice Kennedy Shriver National Institute of Child Health and Human Development (NICHD) Pediatric Terminology ^25^. Severe disease was defined as having received non-invasive ventilation, invasive ventilation or admission to the paediatric intensive care unit (PICU) during the hospital admission.

## Statistical analysis

Descriptive statistics were calculated for baseline characteristics. Continuous variables were summarised as median (with interquartile range) and categorical variables as frequency (percentage). The chi-squared test or Fisher’s exact test was used for testing hypotheses on differences in proportions between groups. The Wilcoxon rank-sum test was used for testing the hypotheses on differences between groups.

We performed multivariable logistic regression to investigate associations of demographic characteristics, co-morbidities (limited to those reported in ≥5% of participants), presence of pneumonia during acute infection and severity of Covid-19 with persistent symptom categories presence at the time of the follow-up interview. We included all participants for whom the variables of interest were available in the final analysis, without imputing missing data. The differing denominators used indicate missing data. Odds ratios were calculated together with 95% confidence intervals (CIs).

Upset plots were used to present the coexistence of persistent symptom categories. Two-sided p-values were reported for all statistical tests, a p-value below 0.05 was considered to be statistically significant. Statistical analysis was performed using R version 3.5.1. Packages used included dplyr, lubridate, ggplots2, plotrix and UpSetR.

### Patient and public involvement

The survey was developed by the ISARIC Global Paediatric Covid-19 follow up working group and informed by a wide range of global stakeholders with expertise in infectious diseases, critical care, paediatrics, epidemiology, allergy-immunology, respiratory medicine, psychiatry, psychology and methodology and patient representatives. The survey was distributed to the members of the patient group and suggestions from parents/carers were implemented.

## RESULTS

### Study population

All 853 children hospitalised with suspected Covid-19 to the hospital between April 2, 2020 and August 26, 2020 were discharged alive (**Figure 1**). Of 836 patients with accurate contact information, parents of 518 RT-PCR positive children agreed to be interviewed (response rate 62%) and were included in the analysis.

**Figure 1.**
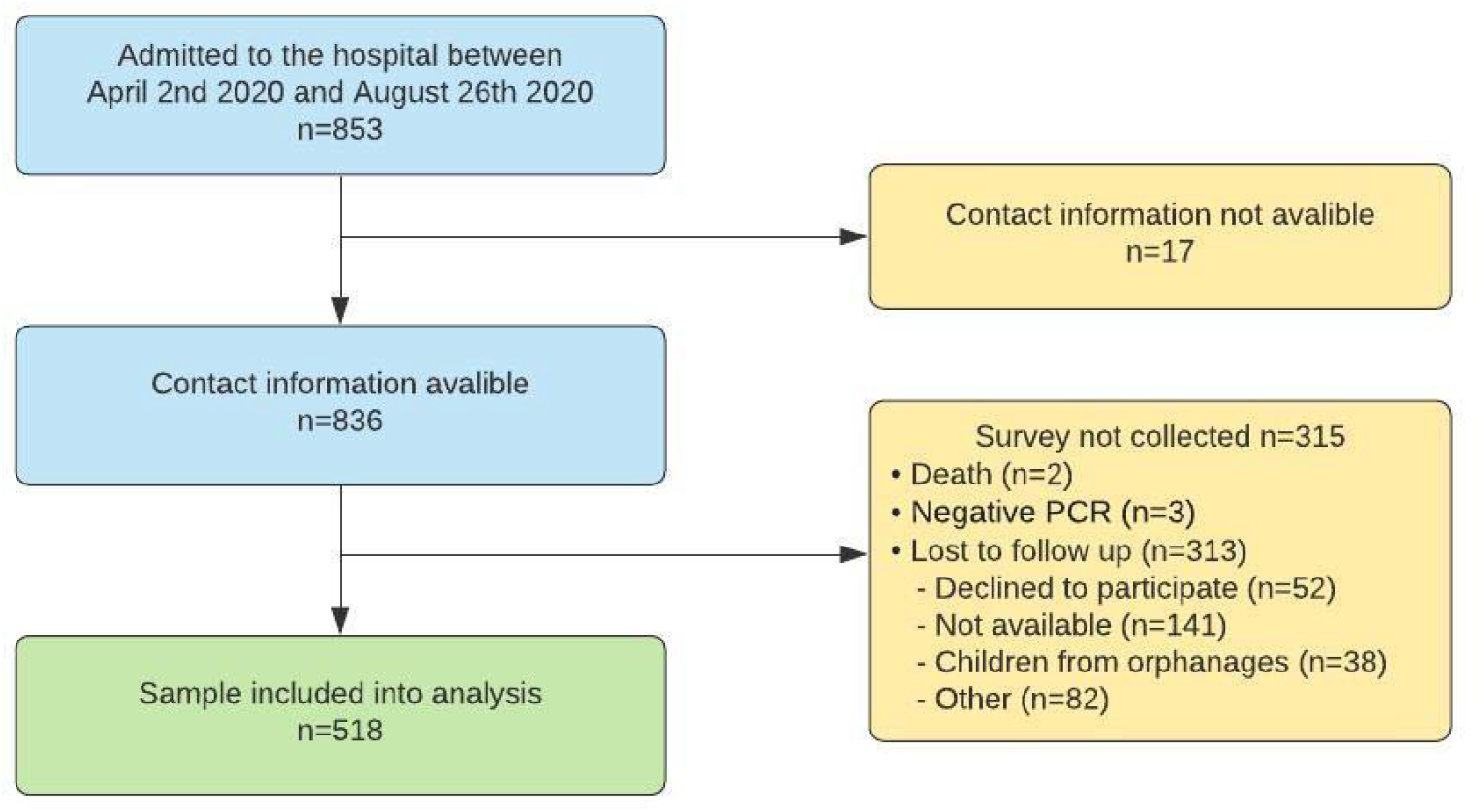
Flow diagram of patients with COVID-19 admitted to Z.A. Bashlyaeva Children’s Municipal Clinical Hospital between April 2, 2020 and August 26, 2020.

The median age was 10.4 years (IQR, 3-15.2; range, 2 days–18 years), 272 (52.2%) were girls. Median follow-up time since hospital admission was 268 days (IQR 233-284). Children had a median of 8 (IQR, 4-9) years of formal school education and a median of 4 (IQR, 3-5) family members were residing in the household (**Table 1**).

**Table 1.** Demographic characteristics of patients admitted to the Z.A. Bashlyaeva Children’s Municipal Clinical Hospital. Data are n (%) or median (IQR) excluding missing values. *All cases of diabetes were type 1. †Allergic diseases include asthma, allergic rhinitis, food allergy and eczema.

| Characteristics | Total (n=518) |
| --- | --- |
| Sex, female | 270 (52.1%) |
| Age at the time of hospital admission, years (median, IQR) | 10.4 (3-15.2) |
| **Age (categorical), years** |  |
| <2 | 105 (20.3%) |
| 2 – 5 | 80 (15.4%) |
| 6 – 11 | 113 (21.8%) |
| 12 – 18 | 220 (42.5%) |
| Days from discharge to follow-up (median, IQR) | 256 (223-271) |
| Length of hospital admission (days, median, IQR) | 10 (7-14) |
| Number of years of formal school education (median, IQR) | 8 (4-9) |
| Number of members in household (median, IQR) | 4 (3-5) |
| Pneumonia during hospitalisation | 192/515 (37.3%) |
| Severe disease (non-invasive ventilation or invasive ventilation or PICU) | 14/515 (2.7%) |
| **Comorbidities** |  |
| Neurological disorders | 43/514 (8.4%) |
| Neurodisability | 11/514 (2.1%) |
| Heart diseases | 21/514 (4.1%) |
| Haematological conditions | 10/514 (1.9%) |
| Tuberculosis | 9/514 (1.8%) |
| Respiratory diseases (not including asthma) | 16/514 (3.1%) |
| Allergic diseases<sup>†</sup> | 121/514 (23.5%) |
| Food Allergy | 67/514 (13%) |
| Allergic Rhinitis | 46/514 (8.9%) |
| Eczema | 45/514 (8.8%) |
| Asthma (doctor diagnosed) | 12/514 (2.3%) |
| Other skin problems (not including eczema) | 8/514 (1.6%) |
| Gastrointestinal problems | 48/514 (9.3%) |
| Oncological conditions | 3/514 (0.6%) |
| Immune system diseases | 6/514 (1.2%) |
| Genetic conditions | 6/514 (1.2%) |
| Diabetes* | 3/514 (0.6%) |
| Other endocrine illness (not diabetes) | 12/514 (2.3%) |
| Renal/Kidney problems | 18/514 (3.5%) |
| Excessive weight and obesity | 25/514 (4.9%) |
| Malnutrition | 10/514 (1.9%) |
| Rheumatological conditions | 4/514 (0.8%) |
| Depression | 4/514 (0.8%) |
| Anxiety | 5/514 (1%) |
| HIV | 0 (0%) |
| No comorbidities | 284/514 (55.3%) |
| One comorbidity | 141/514 (27.4%) |
| Two comorbidities or more | 89/514 (17.3%) |

The most common pre-existing comorbidity in this cohort was food allergy (13%, 67/514), followed by allergic rhinitis and asthma (9.7%, 50/514), gastrointestinal problems (9.3%, 48/514), eczema (8.8%, 45/514) and neurological problems (8.4%, 43/514). Parents of 55.3% (284/514) children did not report any comorbidities. Fever (83.6%, 427/511), cough (55.7%, 284/510), rhinorrhea (54.3%, 278/512) and fatigue (38.9%, 197/506) were the most common presenting symptoms at the time of the hospital admission (**Table S2**). 37.3%, 192/515 of patients had pneumonia during hospital stay, 2.7%, 14/515 had severe disease, which required non-invasive ventilation/invasive ventilation or admission to PICU. Treatments received during the hospital admission are presented in **Table S3**.

At the time of the follow-up interview, parents of 24.7% (128) children reported at least one persistent symptom, with fatigue 10.6% (53/496), insomnia 5.19% (26/501), disturbed smell 4.7% (22/467) and headache 3.5% (17/486) being the most common. Detailed information on symptoms and duration is presented in **Table S4**.

The prevalence of most symptoms declined over time. Prevalence of reported fatigue fell from 15.8% (82/518) at the time of discharge to 11.1% (55/496) 6-7 months later, altered sense of smell fell from 8.7% (45/518) to 5.4% (27/496), altered sense of taste from 5.6% (29/518) to 3.8% (19/496) and breathing difficulties from 3.9% (20/518) to 1.4% (7/496), respectively. However, no change was observed in the prevalence of sleep disturbances or headache over time. Changes in the prevalence of the most common symptoms from the time of discharge over the next 6-7 months are shown in **Figure 2**.

**Figure 2.**
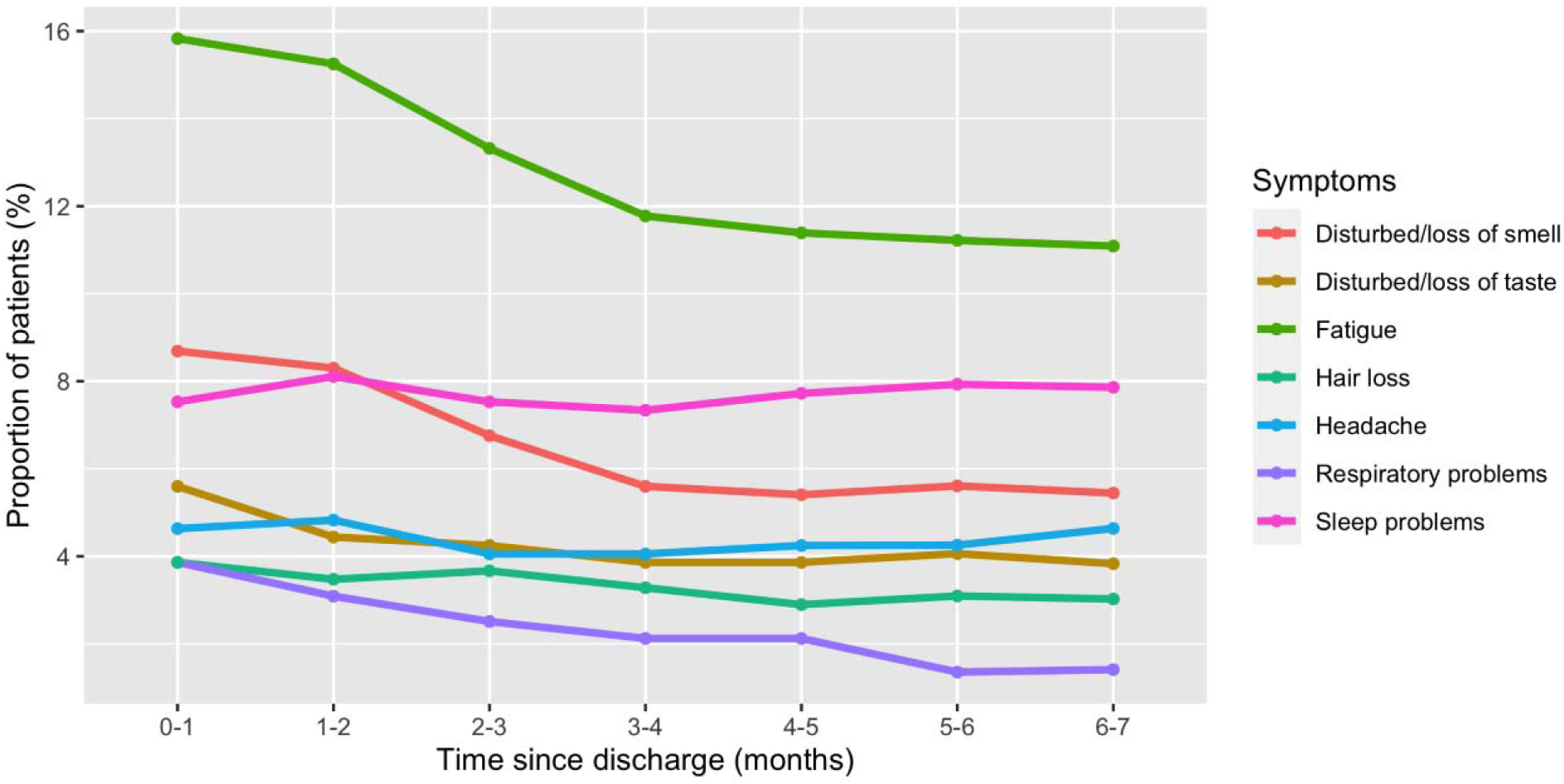
Prevalence of the most common symptoms after hospital discharge. The prevalence was calculated based on responses to the following questions: “Within the last seven days, has your child had any of these symptoms, which were NOT present prior to their Covid-19 illness? (If yes, please indicate below and the duration of the symptom/s) and “Please report any symptoms that have been bothering your child since discharge that are not present today. Please specify the time of onset and duration of these symptoms.”

With regard to persistent symptom categories (**Table S1**), fatigue was the most commonly reported in 10.6% (53/498) of patients at the time of assessment, followed by sleep disturbance 7.2% (36/501), sensory problems 6.2% (29/467), gastrointestinal 4.4% (22/499) and dermatological 3.6% (18/496) problems. A smaller number of patients experienced neurological 3% (14/465), respiratory 2.5% (12/489), cardiovascular 1.9% (9/470) and musculoskeletal 1.8% (9/489) problems long-term.

A total of 8.5% (44) participants reported persistent symptoms from more than one category at the time of the follow-up assessment. Most commonly co-occurring categories were fatigue and sleep problems in 1.9% (10) of children, and fatigue and sensory problems were present in 1.5% (8) of participants. 2.7% (14) of children had persistent symptoms from three or more different categories. Co-existence of persistent symptom categories at the time of the follow-up is presented in the upset plot (**Figure 3**).

**Figure 3.**
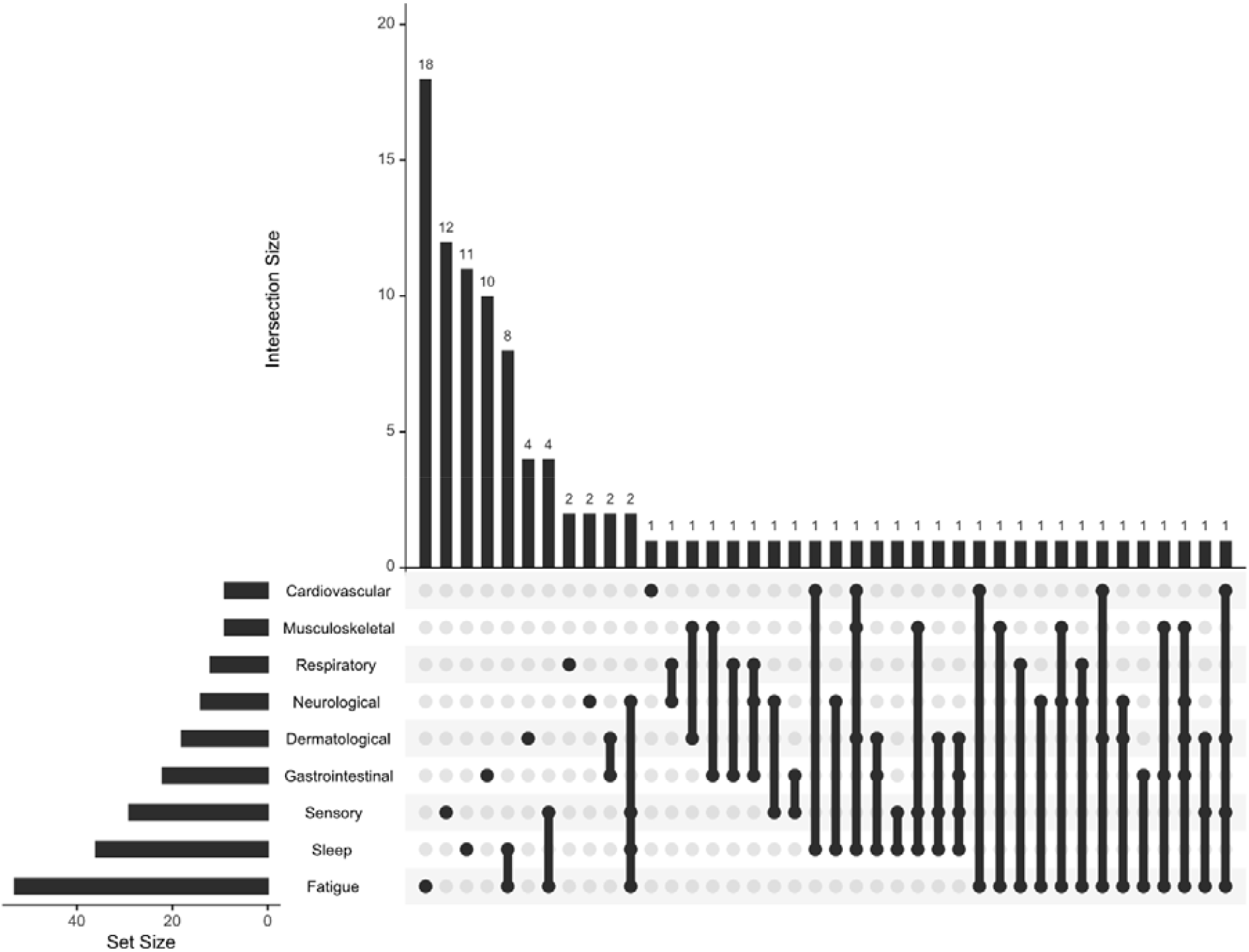
Upset plot representing coexistence of the persistent symptom (present at the time of the follow-up interview and lasting for over 5 months) categories at the follow-up assessment. The values represent the number of individuals experiencing a persistent symptom category or combination of categories. Black lines link multiple symptoms indicated by black dots.

**Figure 4.**
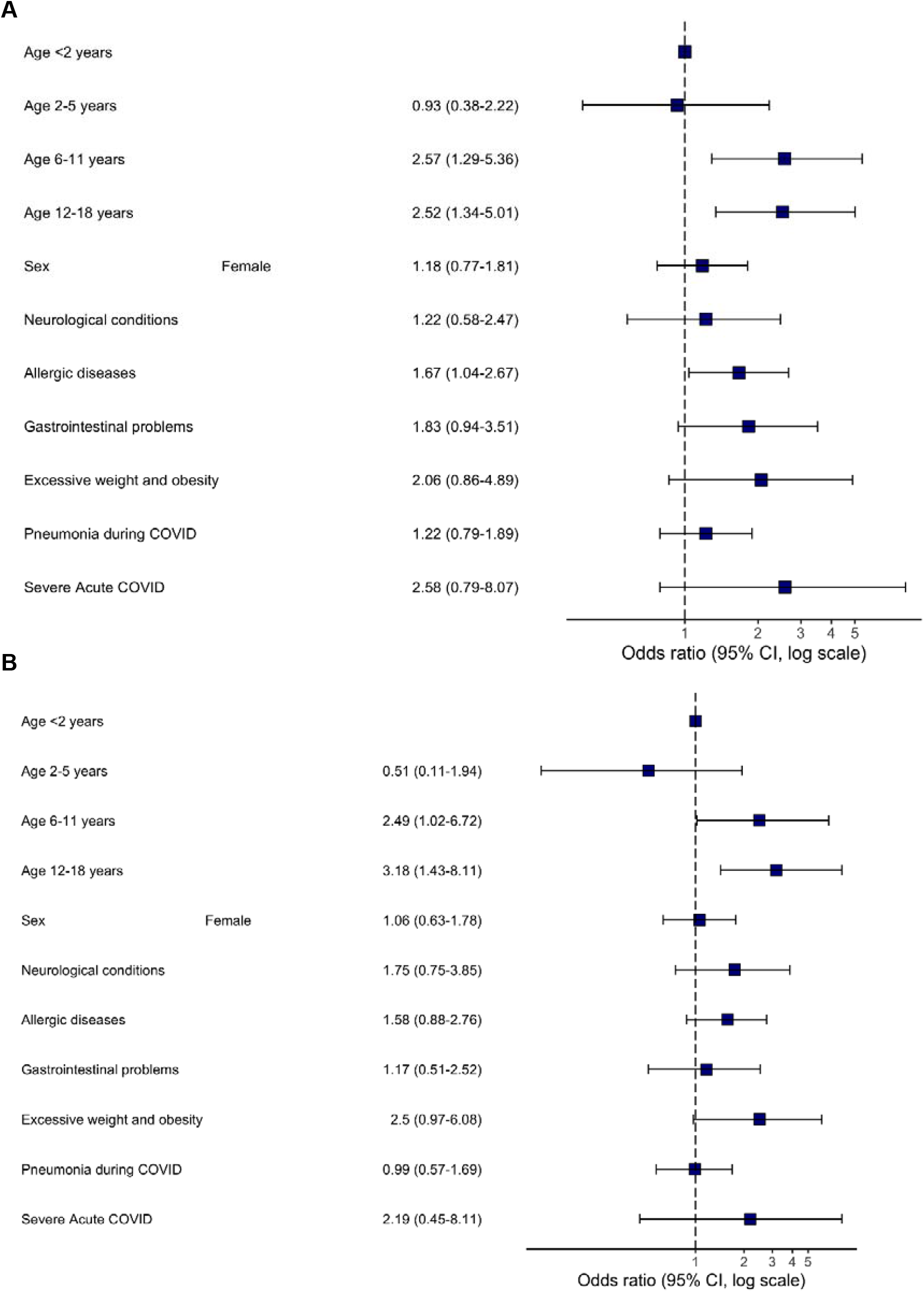
Multivariable logistic regression model to identify pre-existing risk factors for post-COVID condition. Odds ratios and 95% CIs for presence of (A) any category of persistent symptoms at the time of follow-up and (B) two or more co-existing categories of persistent symptoms at the time of the follow-up. Abbreviation: CI, confidence interval.

The scores on the wellness scale for children with one or two or more persistent symptoms significantly declined when compared to before Covid-19 onset from 90 (80-100) to 82.5 (70-93.8) and from 90 (80-95) to 70 (60-80) (p<0.001 for all comparisons), respectively. Children who did not experience any persistent symptoms did not report any significant changes in wellness when asked to compare to how they felt before their acute Covid-19 illness. We also assessed emotional difficulties, social relationships, and activity levels in children (**Tables S4 - S5**). Parents related the following changes to Covid-19 illness, and not to the pandemic in general: less eating in 4.5% (23/512) of children, less sleeping in 3.5% (18/511) and more sleeping in 2% (10/511), reduced physical activity in 4.7% (24/512) and child becoming less emotional in 4.3% (22/511). In contrast, parents attributed changes to social activities to the pandemic in general rather than to the Covid-19 illness: 12% (58/485) of children were spending less time with their friends in person, while 13% (61/470) were spending more time with friends remotely, with less than one percent of parents attributing these changes to Covid-19 illness. 23% (110/478) of children were spending more time watching television, playing video/computer games or using social media for educational purposes, with 92.9% of parents associating these changes with the pandemic in general rather than the Covid-19 illness.

In multivariable regression analysis, older age group was associated with persistent symptoms. When compared with children under two years of ages, those ages 6-11 years had an odds ratio of 2.74 (95% confidence interval 1.37 to 5.75) of persistent symptoms and those 12-18 years of age (OR 2.68, 95% CI 1.41 to 5.4) both vs. <2 years. Another predictor associated with persistent symptoms was allergic diseases (OR 1.67, 95% CI 1.04 to 2.67). Similar patterns were seen for children with co-existence of persistent symptoms from 2 or more categories: 6-11 years of age (OR 2.49, 95% CI 1.02 to 6.72), 12-18 years of age (OR 3.18, 95% CI 1.43 to 8.11) both vs. <2 years.

## DISCUSSION

To our knowledge, this is the largest prospective paediatric cohort study with the longest follow-up, assessing symptom prevalence and duration of long COVID in children and adolescents with laboratory confirmed SARS-CoV-2 infection post hospital discharge. We found that a quarter of children and adolescents had persistent symptoms at the time of the follow-up with fatigue, sleep disturbance and sensory problems being the most common. Almost one in ten reported multi-system impacts with two or more categories of persistent symptoms at the time of the follow-up. Children in mid-childhood and adolescence (age 6-18) were at higher risk of persistent symptoms at the time of the follow-up. Although prevalence of some symptoms declined over time, a substantial proportion experienced problems up to 7-8 months after discharge.

There are very few studies assessing long COVID in children and adolescents; a previous smaller study from Italy found similar persisting symptoms during a shorter follow-up ^26^. It is worth noting that although many parents reported nasal congestion/rhinorrhea in both studies, we found that nasal symptoms persisted long-term in a very small number of children. This highlights importance of symptom duration assessment as point prevalence of the symptom may not reflect its persistence and relevance to requirements for medical care and health/economics.

Although many children experienced symptoms, such as fatigue, disturbed smell and taste, sleep and respiratory problems, hair loss and headaches at the time of the hospital discharge, we witnessed a steady decline in the symptom prevalence over time. This was particularly evident for fatigue and smell disturbance. Prevalence of some symptoms such as headache, and sleep problems did not decline over time, which may be driven by psychological mechanisms rather than pathophysiologic virus infection effects ^27^. A limitation of these findings is that symptom onset and duration was recalled at the single follow-up interview in our study; this may be overcome with repeated follow-ups at appropriate intervals to limit potential recall imprecision. In line with our results, previous research demonstrated symptoms fading over time in adults ^22^ but data are still limited as most of the published cohort studies do not measure symptom duration, but rather assess their presence at a single follow-up.

We found that almost one in ten children had multisystem impacts with two or more categories of persistent symptoms present at the time of the follow-up. Similar numbers were previously reported in the Russian adult population ^3^ and patients with clusters of different symptoms were described in the UK ^28^. Patients with multisystem involvement will represent the primary target for the future research and intervention strategies development.

Age was significantly associated with persistent symptom presence at the time of the follow-up, with children above 6 years of age being at higher risk. To our knowledge, risk factors for long Covid in children have not been investigated in previous studies, so we may draw comparisons with the data from adult cohorts only. Previous data suggest that long Covid is prevalent in adults ^2,3,28-31^ and that age is associated with a higher risk of long Covid ^28,30^. An Australian follow-up study of 151 children aged 0-19 years (median 3 years) who had predominantly mild acute covid-19 infection ^15^ found only 8% with on-going long-covid symptoms. As acknowledged by the authors the low median age may be the main reason for the low long-covid prevalence and our study substantiates this.

We found that allergic diseases in children were also associated with a higher risk of long Covid. This is in agreement with adult studies from Russia ^3^ and the UK ^28^ reporting asthma to be associated with development of long Covid. Recent data suggested that COVID-19 consequences may be linked with the mast cell activation syndrome ^32^ and the Th-2 biased immunological response in children with allergic diseases may be responsible for an increased risk of long-term consequences from the infection. This highlights importance of further research of potential underlying immunological mechanisms of long Covid.

Apart from physical symptoms we assessed emotional and behavioural changes. Although most parents reported no changes, one in twenty parents noticed changes in their children, which they attributed to Covid-19 illness rather than the general situation during the pandemic. These included changes in eating, sleeping, emotional wellbeing and physical activities. Over one in ten parents noted that their children were spending less time in face-to-face communication and more time interacting with their friends remotely and spending time online for both educational and non-educational purposes. These changes were largely attributed to the general situation during the pandemic rather than to the Covid-19 illness. Of note, all surveyed children in our study had PCR-confirmed Covid-19 illness. It is possible, however, that the predominance in remote vs in person interactions will have longer term impacts in themselves. At present, to our knowledge, all publications on long Covid are uncontrolled cohorts due to the difficulties of ascertaining data among controls matched for age and sex but most importantly matched for the same experiences during the pandemic aside from confirmed Covid-19 illness. There is a need to conduct studies with reference groups of non-infected children to reliably assess mental health effects of the pandemic and distinguish them from Covid-19 infection consequences.

A major strength of this study is that it was based on the ISARIC COVID-19 Health and Wellbeing Follow Up Survey for Children which will assist with data harmonisation and comparison with other international studies in the future. Another strength is the large sample size of confirmed Covid-19 infected children, and this cohort has the longest follow-up assessment of hospitalised children to date. Stratification to determine if the symptoms were persistent following Covid-19 and assessment of trends over time were other novel aspects of the study. At the same time, this cohort study has several limitations. First, the study population only included patients within Moscow, although regional clustering is common to many cohort studies published during the Covid-19 pandemic. Second, it included only hospitalised children, not representative of paediatric population. Third, we did not have a control group of previously hospitalised children not experiencing Covid-19 infection. Fourth, some patients may have developed additional comorbidities or complications since the hospital discharge, which were not appropriately captured and could potentially affect the wellbeing and symptom prevalence and persistence. Fifth, the parents/caregivers were interviewed in this study and not children themselves. There is also a risk of recall bias in reporting symptoms which were non-existent at the time of the follow-up and potential selection bias with those with symptoms more likely to agree to survey.

Our findings have implications for further research. Longer follow up duration and repeated assessments combined with controls and sampling for further studies into the pathophysiology and immunology of post-Covid-19 illness sequelae are needed to inform case definitions, and intervention trials aimed to improve long term outcomes.

Although many symptoms which were present at discharge diminished over time, even eight months after hospital discharge many children experienced persistent symptoms, with fatigue, sensory changes and sleep problems being the most common sequelae. One in ten children experienced multi-system involvement at the time of the follow-up. Age and allergic disease were the main risk factors for persistent symptoms. Future work should to be multidisciplinary, prospective, with a control cohort, repeated sampling and with an ability for children to report their health and wellbeing themselves, accompanied by biological sample collection to establish causative mechanisms for a better understanding of Covid-19 sequelae and help with the phenotype/endotype categorisation. Investigation of immunological aspects of the association between allergic diseases and long-covid development may identify mechanisms and therapeutic targets for management to mitigate potential adverse consequences.

## Data Availability

The data that support the findings of this study are available from the corresponding author, DM, upon reasonable request.

## Acknowledgments

We are very grateful to the Z.A. Bashlyaeva Children’s Municipal Clinical Hospital clinical staff and to the patients, parents, carers and families for their kindness and understanding during these difficult times of COVID-19 pandemic. We would like to express our very great appreciation to ISARIC Global COVID-19 follow-up working group for the survey development. We would like to thank Mr Maksim Kholopov for providing technical support in data collection and database administration. We are very thankful to Eat & Talk, Luch, Black Market and Academia for providing us the workspace in time of need and their support of Covid-19 research. Finally, we would like to extend our gratitude to the Global ISARIC team, the ISARIC global adult and paediatric Covid-19 follow up working group, and ISARIC Global support centre for their continuous support, expertise and for the development of the outbreak ready standardised protocols for the data collection.

