## Supplementary material for "Risk factors for long covid in previously hospitalised children using the ISARIC Global follow-up protocol: A prospective cohort study"

### **Case Report Form (CRF) of Initial Survey: First Follow-Up Time Point**

**The questions were answered by the:**

□ Mother/female caregiver □ Father/male caregiver

**Date you completed the survey (DD/MM/YYYY):** [_D_][_D_]/[_M_][_M_]/[_2_][_0_][_Y_][_Y_]

**What is your child’s date of birth (DD/MM/YYYY):** [_D_][_D_]/[_M_][_M_]/[_Y_][_Y_][_Y_][_Y_]

**1.** **About your child**

**Sex/Gender:** □ Male □ Female □ Prefer not to say

**What is your child’s estimated height (cm):** _______________ □ Not sure

**What is your child’s current estimated weight (kg):** _________________□ Not sure

**What was your child's estimated weight before Covid19 illness (kg):** □ Not sure

**How many other members regularly live in your household, including yourself:** [_Number_]

**Does your child study in school/college/university?** □ Yes □ No

**How many years formal school education has your child had?* [_Number_]**

****including primary school (e.g. from around 6 years depending on country)***

**Does your child study in kindergarten?** □ Yes □ No

**2.** **About your child’s Covid-19 illness - all the questions relate to his/her health and wellbeing)**

**Approximately, what day did you first notice your child was experiencing symptoms of**

**Covid-19?** [_D_][_D_]/[_M_][_M_]/[_2_][_0_][_Y_][_Y_]

**How was your child diagnosed with Covid-19?**

□ Laboratory confirmed (positive PCR, antigen or Antibody test) □ Physician confirmed □ Test result is uncertain

□ Not sure

**Estimated date of your child’s most recent positive SARS-CoV-2 /Covid-19 test:** [_D_][_D_]/[_M_][_M_]/[_2_][_0_][_2_][_Y_]

Indicate if □PCR test □ Antibody test □ Unknown

**Has your child been admitted to hospital due to Covid-19?**	□ Yes	□ No

*(If the answer is “no”, please, move on to the section “3”; if the answer is “yes”, please, proceed with the*

*following questions)*

- **Roughly at what date was your child first admitted to hospital?** [_D_][_D_]/[_M_][_M_]/[_2_][_0_][_Y_][_Y_]
- **Roughly at what date was your child first discharged from hospital?** [_D_][_D_]/[_M_][_M_]/[_2_][_0_][_Y_][_Y_]
- **If yes did they spent any time in the Paediatric Intensive Care Unit (PICU)?** □ Yes □ No □ Not sure
- **Has your child been admitted to hospital after the first acute Covid-19 illness?** □ Yes □ No

If yes, how many times: [_Number_]

Name of hospital/hospitals:

If yes, specify reason/reasons:

**3. About your child’s emotional wellbeing, social relationships and activities’**

To answer the following questions, please **mark an X** on the lines below that shows your opinion on the question.

**A. Compared to before your child’s Covid-19 infection, how much is he/she now doing/experiencing the following**

***If there are changes, please indicate whether you think these are due to the illness itself or to the Covid-19 pandemic***

| ***Eating*** | ***Sleeping*** | |
| --- | --- | --- |
| 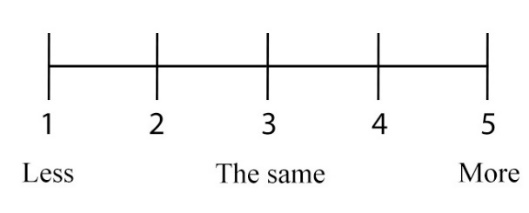  □ Not known | 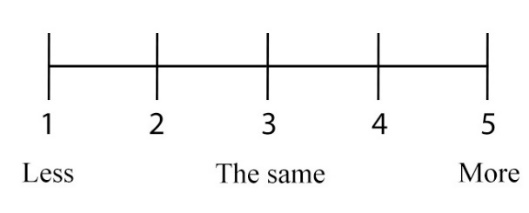  □ Not known | |
| ***If there are changes,*** *please indicate whether you think these are due to*  *□ Illness itself □ Covid-19 pandemic □ Both □ Unsure* | ***If there are changes,*** *please indicate whether you think these are due to*  *□ Illness itself □ Covid-19 pandemic □ Both □ Unsure* | |
| ***Physical Activity*** | | ***Fatigue*** |
| 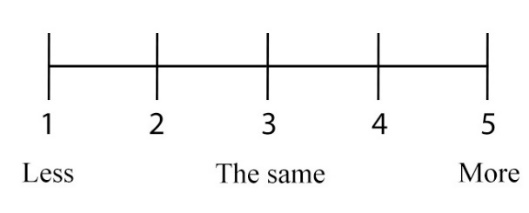  □ Not known | | 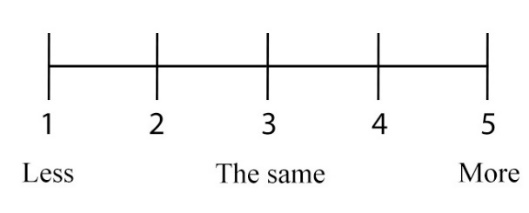  □ Not known |
| ***If there are changes,*** *please indicate whether you think these are due to*  *□ Illness itself □ Covid-19 pandemic □ Both □ Unsure* | | ***If there are changes,*** *please indicate whether you think*  *these are due to*  *□ Illness itself □ Covid-19 pandemic □ Both □ Unsure* |
| ***Spending time with friends in-person*** | | ***Spending time with friends remotely***  ***(e.g., online, social media, texting)*** |
| 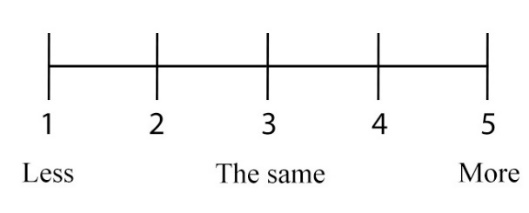  □ Not known | | 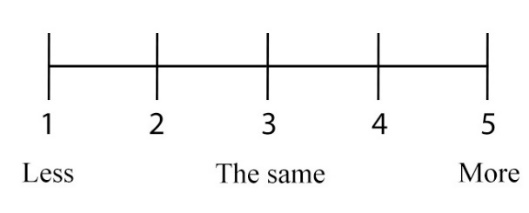  □ Not known |
| ***If there are changes,*** *please indicate whether you think these are due to*  *□ Illness itself □ Covid-19 pandemic □ Both □ Unsure* | | ***If there are changes,*** *please indicate whether you think*  *these are due to*  *□ Illness itself □ Covid-19 pandemic □ Both □ Unsure* |
| ***Spending time watching TV, playing***  ***video/computer games, or using***  ***social media for educational purposes, including school/nursery work*** | | ***Spending time watching TV, playing***  ***video/computer games, or using***  ***social media for non-educational purposes,*** |
| 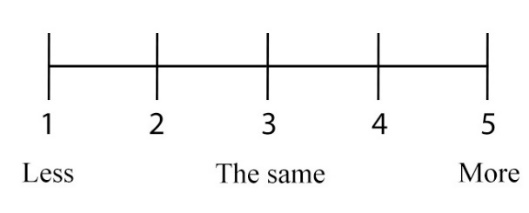  □ Not known | | 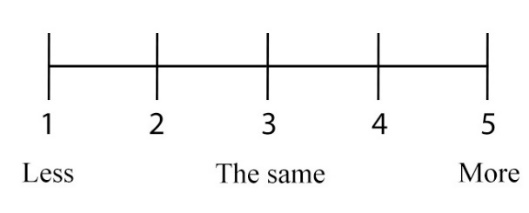  □ Not known |
| ***If there are changes,*** *please indicate whether you think these are due to*  *□ Illness itself □ Covid-19 pandemic □ Both □ Unsure* | | ***If there are changes,*** *please indicate whether you think*  *these are due to*  *□ Illness itself □ Covid-19 pandemic □ Both □ Unsure* |
| ***Spending time outside*** | | ***Attending school/nursery*** |
| 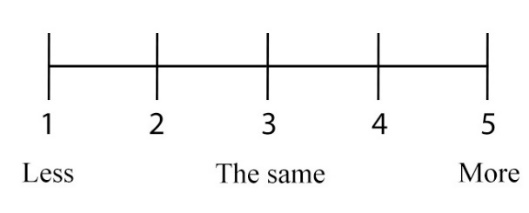  □ Not known | | 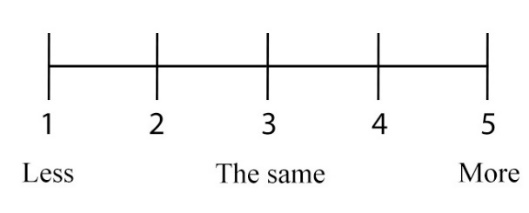  **□** My child has not been attending school/nursery before  Covid-19 infection |
| ***If there are changes,*** *please indicate whether you think these are due to*  *□ Illness itself □ Covid-19 pandemic □ Both □ Unsure* | | ***If there are changes,*** *please indicate whether you think*  *these are due to*  *□ Illness itself □ Covid-19 pandemic □ Both □ Unsure* |

**B. Compared to before your child’s Covid-19 illness: Have there been changes in your child’s…**

| **… CONNECTEDNESS with others…** | **…EMOTIONS?** |
| --- | --- |
| 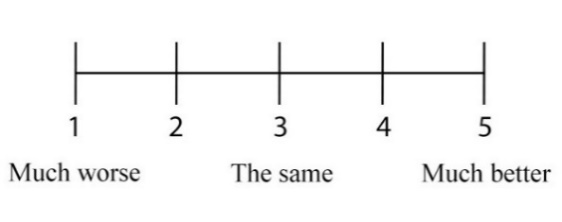  □ Unsure | 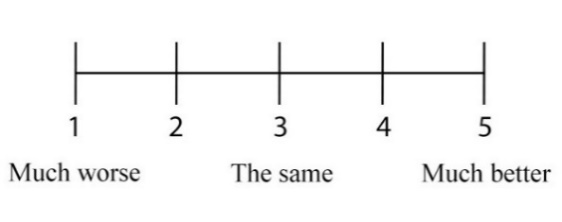  □ Unsure |
| ***If there are changes,*** *please indicate whether you think these are due to*  *□ Illness itself □ Covid-19 pandemic □ Both □Unsure* | ***If there are changes,*** *please indicate whether you think these are due to*  *□ Illness itself □ Covid-19 pandemic □ Both □Unsure* |
| **…BEHAVIOUR?** | **…RELATIONSHIPS, in how they get on with others?** |
| 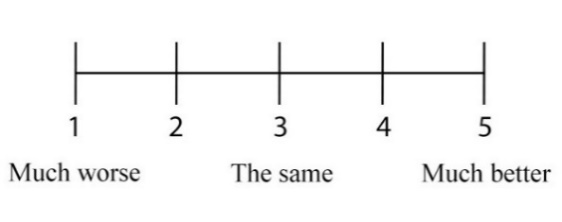  □ Unsure | 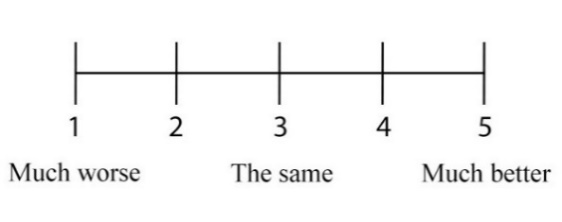  □ Unsure |
| ***If there are changes,*** *please indicate whether you think these are due to*  *□ Illness itself □ Covid-19 pandemic □ Both □Unsure* | ***If there are changes,*** *please indicate whether you think these are due to*  *□ Illness itself □ Covid-19 pandemic □ Both □Unsure* |

C. **Have you asked for help from a health professional because of Covid-19 illness consequences to your child’s EMOTIONS, BEHAVIOUR OR RELATIONSHIPS?**

□ Yes □ No	If yes who did you ask for help from_______________________________

D. **If you have replied MUCH WORSE to any of the options in question “B” OR YOU HAVE ASKED FOR**

**HELP for these problems (question “C”), please, answer 3 of the following questions:**

1. **Do the difficulties upset or distress your child?**

□ Not at all □ Only a little □ Undecided □ Quite a lot □ A great deal

2. **Do the difficulties interfere with your child's everyday life in the following areas?**

| Home Life | □ Not at all □ Only a little □ Undecided □ Quite a lot □ A great deal |
| --- | --- |
| Friendships | □ Not at all □ Only a little □ Undecided □ Quite a lot □ A great deal |
| Classroom Learning | □ Not at all □ Only a little □ Undecided □ Quite a lot □ A great deal |
| Leisure Activities | □ Not at all □ Only a little □ Undecided □ Quite a lot □ A great deal |

3. **Do these difficulties put a burden on you or the family as a whole?**

□ Not at all □ Only a little □ Undecided □ Quite a lot □ A great deal □ Unsure

E. **Do you live in Moscow or Moscow Oblast?** □ Yes □ No

**If No,what is the current situation in your town/city/region on lockdown measures?
*(you may select more than one answer)***

| □ Closing of child’s school | □ Closing of nurseries/kindergartens |
| --- | --- |
| □ Closing of non-essential shops (shops and stores apart from food, doctors and drug stores) | □ Cancellation/closing of recreational venues and activities |
| □ Closing of indoor places/venues | □ Closing of outdoor recreational places |
| □ Constraining meeting friends | □ Stay-at-home orders (not allowed to leave the house except for essential errands) |

**4a. About your child’s state of health prior to his/her Covid-19 illness**

**Has your child been physician’s diagnosed or received treatment/support for any of the following chronic
medical conditions prior to the Covid-19 infection? (answer with a tick in the box)**

|  | **Yes** | **No** | **Unknown** |
| --- | --- | --- | --- |
| Prematurity *(baby born <37 weeks)* |  |  |  |
| Neurological |  |  |  |
| Neurodisability |  |  |  |
| Heart diseases |  |  |  |
| Respiratory diseases (not including asthma) |  |  |  |
| Tuberculosis |  |  |  |
| Asthma (doctor’s diagnosed) |  |  |  |
| Allergic rhinitis/hay fever |  |  |  |
| Food allergy |  |  |  |
| Atopic dermatitis/Eczema |  |  |  |
| Other skin problems (not including eczema) |  |  |  |
| Gut problems |  |  |  |
| Haematology *(blood diseases)* |  |  |  |
| Oncology *(cancer or other progressively enlarging or spreading tumor)* |  |  |  |
| Immune system diseases (*e.g. primary immune deficiencies*) |  |  |  |
| Genetic conditions |  |  |  |
| Diabetes (if yes indicate type: □ Type 1 □ Type 2) |  |  |  |
| Other endocrine illness (not diabetes) |  |  |  |
| Renal/Kidney problems |  |  |  |
| Excessive weight and obesity |  |  |  |
| Malnutrition *(deficiencies, excesses, or imbalances in a person's*  *intake of energy and/or nutrients)* |  |  |  |
| Rheumatology *(e.g. arthritis, or inflammation of the joints)* |  |  |  |
| Depression |  |  |  |
| Anxiety |  |  |  |
| HIV |  |  |  |
| Other (please indicate) |  | | |

**Has your child ever been under Child and Adolescent Mental Health services before the Covid-19 pandemic? □ Yes □ No □ Not sure**

**Prior to Covid-19 infection, how was your child’s physical health in general?**

□ Very poor □ Poor □ Ok □ Good □ Very good

***If you ticked poor or very poor, please explain:***

**Prior to Covid-19 infection, how would you describe your child’s mental health in general**

□ Very poor □ Poor □ Ok □ Good □ Very good

***If you ticked poor or very poor, please explain:***_________________________________________________________________________________

**Have you requested help because of Covid-19 consequences to your child’s physical health?**

□ Yes □ No □ Not sure

**4b. About your child’s current health**

**Has your child felt feverish recently?** □ Yes □ No □ Not sure

***If yes indicate when they felt feverish (tick all that apply)***

□ Within the last 7 days □ >1-2 weeks □ >2-4 weeks □ >1-2 months □ >2-3 months □ >3-6 months

□ >6 months ago

***If yes, what was the most likely cause of your child’s most recent feverish illness?***

□ Covid-19 □ Other respiratory infection (cough/cold/sore throat) □ TB □ Stomach infection (diarrhea/vomiting) □ Urinary infection □ Other (specify): ___________________________________

□ Unknown □ Prefer not to say

**If Covid-19, what was the estimated date of the most recent positive SARS-CoV-2 /Covid-19 test?**

[_D_][_D_]/[_M_][_M_]/[_2_][_0_][_2_][_Y_]

**Indicate if** □PCR test □ Antibody test □ Unknown

**How much do you agree with the following statement?**

**“My child has fully recovered from Covid-19”**

Please **mark an X** on the line below that shows your opinion on the question as of **TODAY**

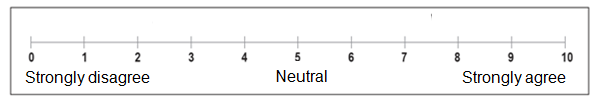

**5. Since having Covid-19, has your child been diagnosed with any of the following? (indicate the correct answer in the box provided)**

|  | **YES** | **NO** |  | **YES** | **NO** |
| --- | --- | --- | --- | --- | --- |
| Multisystem inflammatory syndrome |  |  | Shock / Toxic shock syndrome |  |  |
| Pulmonary embolism  *(PE, “Clot in lung”)* |  |  | Coagulopathy *(excessive bleeding or clotting)* |  |  |
| Kawasaki disease |  |  | Kidney problems |  |  |
| Multisystem inflammatory syndrome (MIS-C/PIMS-TS) |  |  | Type 1 Diabetes |  |  |
| Respiratory failure |  |  | Type 2 Diabetes |  |  |
| Asthma |  |  | Intussusception |  |  |
| Myocarditis  *(inflammation of the heart muscle)* |  |  | Other condition (if yes specify): |  |  |

| **6a. Within the last seven days, has your child had any of these symptoms, which were NOT present prior to their Covid-19 illness?**  **If yes, please indicate below and the duration of the symptom/s:** |
| --- |

| **Respiratory problems** | **Tick Yes or No** | **If yes**, what is the duration of symptoms |  |  |
| --- | --- | --- | --- | --- |
| Nasal congestion / rhinorrhea | □ Yes □ No | □ < 1 month □ 1-2 months □ >2 -3 months □ >3-4 months □ >4-5 months  □ >5-6 months □ >6-7 months □ >7 -8 months □ >8-9 months □ >9 -10 months  □ >10-11 months □ >11 -12 months □ >12 months □ From the time of discharge  □ Not sure |  |  |
| Difficulty breathing /chest tightness | □ Yes □ No | □ < 1 month □ 1-2 months □ >2 -3 months □ >3-4 months □ >4-5 months  □ >5-6 months □ >6-7 months □ >7 -8 months □ >8-9 months □ >9 -10 months  □ >10-11 months □ >11 -12 months □ >12 months □ From the time of discharge  □ Not sure |  |  |
| Pain on breathing | □ Yes □ No | □ < 1 month □ 1-2 months □ >2 -3 months □ >3-4 months □ >4-5 months  □ >5-6 months □ >6-7 months □ >7 -8 months □ >8-9 months □ >9 -10 months  □ >10-11 months □ >11 -12 months □ >12 months □ From the time of discharge  □ Not sure |  |  |
| Chest pain | □ Yes □ No | □ < 1 month □ 1-2 months □ >2 -3 months □ >3-4 months □ >4-5 months  □ >5-6 months □ >6-7 months □ >7 -8 months □ >8-9 months □ >9 -10 months  □ >10-11 months □ >11 -12 months □ >12 months □ From the time of discharge  □ Not sure |  |  |
| Persistent cough | □ Yes □ No | □ < 1 month □ 1-2 months □ >2 -3 months □ >3-4 months □ >4-5 months  □ >5-6 months □ >6-7 months □ >7 -8 months □ >8-9 months □ >9 -10 months  □ >10-11 months □ >11 -12 months □ >12 months □ From the time of discharge  □ Not sure |  |  |
| *If yes, □ dry cough □ with phlegm* | | |  |  |
| **Musculoskeletal problems** | **Tick Yes or No** | **If yes**, what is the duration of symptoms |  |  |
| Cannot fully move or control movement | □ Yes □ No | □ < 1 month □ 1-2 months □ >2 -3 months □ >3-4 months □ >4-5 months  □ >5-6 months □ >6-7 months □ >7 -8 months □ >8-9 months □ >9 -10 months  □ >10-11 months □ >11 -12 months □ >12 months □ From the time of discharge  □ Not sure |  |  |
| Problems with balance | □ Yes □ No | □ < 1 month □ 1-2 months □ >2 -3 months □ >3-4 months □ >4-5 months  □ >5-6 months □ >6-7 months □ >7 -8 months □ >8-9 months □ >9 -10 months  □ >10-11 months □ >11 -12 months □ >12 months □ From the time of discharge  □ Not sure |  |  |
| Persistent muscle pain | □ Yes □ No | □ < 1 month □ 1-2 months □ >2 -3 months □ >3-4 months □ >4-5 months  □ >5-6 months □ >6-7 months □ >7 -8 months □ >8-9 months □ >9 -10 months  □ >10-11 months □ >11 -12 months □ >12 months □ From the time of discharge  □ Not sure |  |  |
| Joint pain or swelling | □ Yes □ No | □ < 1 month □ 1-2 months □ >2 -3 months □ >3-4 months □ >4-5 months  □ >5-6 months □ >6-7 months □ >7 -8 months □ >8-9 months □ >9 -10 months  □ >10-11 months □ >11 -12 months □ >12 months □ From the time of discharge  □ Not sure |  |  |
| **Neurological problems** | **Tick Yes or No** | **If yes**, what is the duration of symptoms |  |  |
| Headache | □ Yes □ No | □ < 1 month □ 1-2 months □ >2 -3 months □ >3-4 months □ >4-5 months  □ >5-6 months □ >6-7 months □ >7 -8 months □ >8-9 months □ >9 -10 months  □ >10-11 months □ >11 -12 months □ >12 months □ From the time of discharge  □ Not sure |  |  |
| Dizziness/ light headedness | □ Yes □ No | □ < 1 month □ 1-2 months □ >2 -3 months □ >3-4 months □ >4-5 months  □ >5-6 months □ >6-7 months □ >7 -8 months □ >8-9 months □ >9 -10 months  □ >10-11 months □ >11 -12 months □ >12 months □ From the time of discharge  □ Not sure |  |  |
| Fainting/ blackouts | □ Yes □ No | □ < 1 month □ 1-2 months □ >2 -3 months □ >3-4 months □ >4-5 months  □ >5-6 months □ >6-7 months □ >7 -8 months □ >8-9 months □ >9 -10 months  □ >10-11 months □ >11 -12 months □ >12 months □ From the time of discharge  □ Not sure |  |  |
| Problems seeing/blurred vision | □ Yes □ No | □ < 1 month □ 1-2 months □ >2 -3 months □ >3-4 months □ >4-5 months  □ >5-6 months □ >6-7 months □ >7 -8 months □ >8-9 months □ >9 -10 months  □ >10-11 months □ >11 -12 months □ >12 months □ From the time of discharge  □ Not sure |  |  |
| Disturbed smell | □ Yes □ No | □ < 1 month □ 1-2 months □ >2 -3 months □ >3-4 months □ >4-5 months  □ >5-6 months □ >6-7 months □ >7 -8 months □ >8-9 months □ >9 -10 months  □ >10-11 months □ >11 -12 months □ >12 months □ From the time of discharge  □ Not sure |  |  |
| Loss of smell | □ Yes □ No | □ < 1 month □ 1-2 months □ >2 -3 months □ >3-4 months □ >4-5 months  □ >5-6 months □ >6-7 months □ >7 -8 months □ >8-9 months □ >9 -10 months  □ >10-11 months □ >11 -12 months □ >12 months □ From the time of discharge  □ Not sure |  |  |
| Disturbed taste | □ Yes □ No | □ < 1 month □ 1-2 months □ >2 -3 months □ >3-4 months □ >4-5 months  □ >5-6 months □ >6-7 months □ >7 -8 months □ >8-9 months □ >9 -10 months  □ >10-11 months □ >11 -12 months □ >12 months □ From the time of discharge  □ Not sure |  |  |
| Loss of taste | □ Yes □ No | □ < 1 month □ 1-2 months □ >2 -3 months □ >3-4 months □ >4-5 months  □ >5-6 months □ >6-7 months □ >7 -8 months □ >8-9 months □ >9 -10 months  □ >10-11 months □ >11 -12 months □ >12 months □ From the time of discharge  □ Not sure |  |  |
| Tremor/shakiness | □ Yes □ No | □ < 1 month □ 1-2 months □ >2 -3 months □ >3-4 months □ >4-5 months  □ >5-6 months □ >6-7 months □ >7 -8 months □ >8-9 months □ >9 -10 months  □ >10-11 months □ >11 -12 months □ >12 months □ From the time of discharge  □ Not sure |  |  |
| Tingling feeling/ “pins and needles“ | □ Yes □ No | □ < 1 month □ 1-2 months □ >2 -3 months □ >3-4 months □ >4-5 months  □ >5-6 months □ >6-7 months □ >7 -8 months □ >8-9 months □ >9 -10 months  □ >10-11 months □ >11 -12 months □ >12 months □ From the time of discharge  □ Not sure |  |  |
| Seizures/fits | □ Yes □ No | □ < 1 month □ 1-2 months □ >2 -3 months □ >3-4 months □ >4-5 months  □ >5-6 months □ >6-7 months □ >7 -8 months □ >8-9 months □ >9 -10 months  □ >10-11 months □ >11 -12 months □ >12 months □ From the time of discharge  □ Not sure |  |  |
| Confusion/lack of concentration | □ Yes □ No | □ < 1 month □ 1-2 months □ >2 -3 months □ >3-4 months □ >4-5 months  □ >5-6 months □ >6-7 months □ >7 -8 months □ >8-9 months □ >9 -10 months  □ >10-11 months □ >11 -12 months □ >12 months □ From the time of discharge  □ Not sure |  |  |
| Problems speaking or communicating | □ Yes □ No | □ < 1 month □ 1-2 months □ >2 -3 months □ >3-4 months □ >4-5 months  □ >5-6 months □ >6-7 months □ >7 -8 months □ >8-9 months □ >9 -10 months  □ >10-11 months □ >11 -12 months □ >12 months □ From the time of discharge  □ Not sure |  |  |
| Insomnia *(hard to fall asleep, hard to stay asleep)* | □ Yes □ No | □ < 1 month □ 1-2 months □ >2 -3 months □ >3-4 months □ >4-5 months  □ >5-6 months □ >6-7 months □ >7 -8 months □ >8-9 months □ >9 -10 months  □ >10-11 months □ >11 -12 months □ >12 months □ From the time of discharge  □ Not sure |  |  |
| Hypersomnia *(excessive daytime sleepiness or prolonged nighttime sleep)* | □ Yes □ No | □ < 1 month □ 1-2 months □ >2 -3 months □ >3-4 months □ >4-5 months  □ >5-6 months □ >6-7 months □ >7 -8 months □ >8-9 months □ >9 -10 months  □ >10-11 months □ >11 -12 months □ >12 months □ From the time of discharge  □ Not sure |  |  |
| **Fatigue** | □ Yes □ No | □ < 1 month □ 1-2 months □ >2 -3 months □ >3-4 months □ >4-5 months  □ >5-6 months □ >6-7 months □ >7 -8 months □ >8-9 months □ >9 -10 months  □ >10-11 months □ >11 -12 months □ >12 months □ From the time of discharge  □ Not sure |  |  |
| **Gastrointestinal problems** | **Tick Yes or No** | **If yes**, what is the duration of symptoms |  |  |
| Weight loss | □ Yes □ No | □ < 1 month □ 1-2 months □ >2 -3 months □ >3-4 months □ >4-5 months  □ >5-6 months □ >6-7 months □ >7 -8 months □ >8-9 months □ >9 -10 months  □ >10-11 months □ >11 -12 months □ >12 months □ From the time of discharge  □ Not sure |  |  |
| Problems swallowing or chewing | □ Yes □ No | □ < 1 month □ 1-2 months □ >2 -3 months □ >3-4 months □ >4-5 months  □ >5-6 months □ >6-7 months □ >7 -8 months □ >8-9 months □ >9 -10 months  □ >10-11 months □ >11 -12 months □ >12 months □ From the time of discharge  □ Not sure |  |  |
| Poor appetite | □ Yes □ No | □ < 1 month □ 1-2 months □ >2 -3 months □ >3-4 months □ >4-5 months  □ >5-6 months □ >6-7 months □ >7 -8 months □ >8-9 months □ >9 -10 months  □ >10-11 months □ >11 -12 months □ >12 months □ From the time of discharge  □ Not sure |  |  |
| Diarrhea | □ Yes □ No | □ < 1 month □ 1-2 months □ >2 -3 months □ >3-4 months □ >4-5 months  □ >5-6 months □ >6-7 months □ >7 -8 months □ >8-9 months □ >9 -10 months  □ >10-11 months □ >11 -12 months □ >12 months □ From the time of discharge  □ Not sure |  |  |
| Stomach/ abdominal pain | □ Yes □ No | □ < 1 month □ 1-2 months □ >2 -3 months □ >3-4 months □ >4-5 months  □ >5-6 months □ >6-7 months □ >7 -8 months □ >8-9 months □ >9 -10 months  □ >10-11 months □ >11 -12 months □ >12 months □ From the time of discharge  □ Not sure |  |  |
| Feeling nauseous | □ Yes □ No | □ < 1 month □ 1-2 months □ >2 -3 months □ >3-4 months □ >4-5 months  □ >5-6 months □ >6-7 months □ >7 -8 months □ >8-9 months □ >9 -10 months  □ >10-11 months □ >11 -12 months □ >12 months □ From the time of discharge  □ Not sure |  |  |
| Vomiting | □ Yes □ No | □ < 1 month □ 1-2 months □ >2 -3 months □ >3-4 months □ >4-5 months  □ >5-6 months □ >6-7 months □ >7 -8 months □ >8-9 months □ >9 -10 months  □ >10-11 months □ >11 -12 months □ >12 months □ From the time of discharge  □ Not sure |  |  |
| Constipation | □ Yes □ No | □ < 1 month □ 1-2 months □ >2 -3 months □ >3-4 months □ >4-5 months  □ >5-6 months □ >6-7 months □ >7 -8 months □ >8-9 months □ >9 -10 months  □ >10-11 months □ >11 -12 months □ >12 months □ From the time of discharge  □ Not sure |  |  |
| **Cardiovascular problems** | **Tick Yes or No** | **If yes**, what is the duration of symptoms |  |  |
| Palpitations (heart racing) | □ Yes □ No | □ < 1 month □ 1-2 months □ >2 -3 months □ >3-4 months □ >4-5 months  □ >5-6 months □ >6-7 months □ >7 -8 months □ >8-9 months □ >9 -10 months  □ >10-11 months □ >11 -12 months □ >12 months □ From the time of discharge  □ Not sure |  |  |
| Variations in heart rate (tachycardia or bradycardia) | □ Yes □ No | □ < 1 month □ 1-2 months □ >2 -3 months □ >3-4 months □ >4-5 months  □ >5-6 months □ >6-7 months □ >7 -8 months □ >8-9 months □ >9 -10 months  □ >10-11 months □ >11 -12 months □ >12 months □ From the time of discharge  □ Not sure |  |  |
| Bleeding | □ Yes □ No | □ < 1 month □ 1-2 months □ >2 -3 months □ >3-4 months □ >4-5 months  □ >5-6 months □ >6-7 months □ >7 -8 months □ >8-9 months □ >9 -10 months  □ >10-11 months □ >11 -12 months □ >12 months □ From the time of discharge  □ Not sure |  |  |
| *If yes, specify bleeding site:* |  |  |  |  |
| **Genitourinary problems** | **Tick Yes or No** | **If yes**, what is the duration of symptoms |  | **If yes**, what is the duration of symptoms |
| Urination problems | □ Yes □ No | □ < 1 month □ 1-2 months □ >2 -3 months □ >3-4 months □ >4-5 months  □ >5-6 months □ >6-7 months □ >7 -8 months □ >8-9 months □ >9 -10 months  □ >10-11 months □ >11 -12 months □ >12 months □ From the time of discharge  □ Not sure |  |  |
| Changes in menstruation, (if regular before Covid-19 illness) | □ Yes □ No  □ Not applicable | □ < 1 month □ 1-2 months □ >2 -3 months □ >3-4 months □ >4-5 months  □ >5-6 months □ >6-7 months □ >7 -8 months □ >8-9 months □ >9 -10 months  □ >10-11 months □ >11 -12 months □ >12 months □ From the time of discharge  □ Not sure |  |  |
| **Other problems** | **Tick Yes or No** | **If yes**, what is the duration of symptoms |  |  |
| Bilateral conjunctivitis  *If yes, □ purulent □ non-purulent* | □ Yes □ No | □ < 1 month □ 1-2 months □ >2 -3 months □ >3-4 months □ >4-5 months  □ >5-6 months □ >6-7 months □ >7 -8 months □ >8-9 months □ >9 -10 months  □ >10-11 months □ >11 -12 months □ >12 months □ From the time of discharge  □ Not sure |  |  |
| Lumps or rashes (purple/pink) on toes | □ Yes □ No | □ < 1 month □ 1-2 months □ >2 -3 months □ >3-4 months □ >4-5 months  □ >5-6 months □ >6-7 months □ >7 -8 months □ >8-9 months □ >9 -10 months  □ >10-11 months □ >11 -12 months □ >12 months □ From the time of discharge  □ Not sure |  |  |
| Skin rash | □ Yes □ No | □ < 1 month □ 1-2 months □ >2 -3 months □ >3-4 months □ >4-5 months  □ >5-6 months □ >6-7 months □ >7 -8 months □ >8-9 months □ >9 -10 months  □ >10-11 months □ >11 -12 months □ >12 months □ From the time of discharge  □ Not sure |  |  |
| Skin rash *If yes, tick all body areas that apply:* | □ Yes □ No | □ Face  □ Trunk (stomach or back)  □ Arms  □ Legs  □ Buttocks  □ Toes  □ Fingers  □ Accompanied by itch |  |  |
| Hair loss | □ Yes □ No | □ < 1 month □ 1-2 months □ >2 -3 months □ >3-4 months □ >4-5 months  □ >5-6 months □ >6-7 months □ >7 -8 months □ >8-9 months □ >9 -10 months  □ >10-11 months □ >11 -12 months □ >12 months □ From the time of discharge  □ Not sure |  |  |
| Hyperhidrosis | □ Yes □ No | □ < 1 month □ 1-2 months □ >2 -3 months □ >3-4 months □ >4-5 months  □ >5-6 months □ >6-7 months □ >7 -8 months □ >8-9 months □ >9 -10 months  □ >10-11 months □ >11 -12 months □ >12 months □ From the time of discharge  □ Not sure |  |  |
| **Other New Symptoms, if yes, specify all with their duration:** | | **If yes**, what is the duration of symptoms |  |  |
|  | | □ < 1 month □ 1-2 months □ >2 -3 months □ >3-4 months □ >4-5 months  □ >5-6 months □ >6-7 months □ >7 -8 months □ >8-9 months □ >9 -10 months  □ >10-11 months □ >11 -12 months □ >12 months □ From the time of discharge  □ Not sure |  |  |
|  | | □ < 1 month □ 1-2 months □ >2 -3 months □ >3-4 months □ >4-5 months  □ >5-6 months □ >6-7 months □ >7 -8 months □ >8-9 months □ >9 -10 months  □ >10-11 months □ >11 -12 months □ >12 months □ From the time of discharge  □ Not sure |  |  |

**6b. Please report any symptoms that have been bothering your child since discharge that are not present today. Please specify the time of onset and duration of these symptoms**

| **Respiratory problems** | **Tick Yes or No** | **If yes**, what was the time of onset |  |  |
| --- | --- | --- | --- | --- |
| Nasal congestion / rhinorrhea | □ Yes □ No | □ < 1 month □ 1-2 months □ >2 -3 months □ >3-4 months □ >4-5 months  □ >5-6 months □ >6-7 months □ >7 -8 months □ >8-9 months □ >9 -10 months  □ >10-11 months □ >11 -12 months □ >12 months □ From the time of discharge  □ Not sure |  |  |
|  |  | **If yes**, what was the duration of symptoms |  |  |
|  |  | □ < 1 month □ 1-2 months □ >2 -3 months □ >3-4 months □ >4-5 months  □ >5-6 months □ >6-7 months □ >7 -8 months □ >8-9 months □ >9 -10 months  □ >10-11 months □ >11 -12 months □ >12 months □ Not sure |  |  |
| Difficulty breathing /chest tightness | □ Yes □ No | **If yes**, what was the time of onset |  |  |
|  |  | □ < 1 month □ 1-2 months □ >2 -3 months □ >3-4 months □ >4-5 months  □ >5-6 months □ >6-7 months □ >7 -8 months □ >8-9 months □ >9 -10 months  □ >10-11 months □ >11 -12 months □ >12 months □ From the time of discharge  □ Not sure |  |  |
|  |  | **If yes**, what was the duration of symptoms |  |  |
|  |  | □ < 1 month □ 1-2 months □ >2 -3 months □ >3-4 months □ >4-5 months  □ >5-6 months □ >6-7 months □ >7 -8 months □ >8-9 months □ >9 -10 months  □ >10-11 months □ >11 -12 months □ >12 months □ Not sure |  |  |
| Pain on breathing | □ Yes □ No | **If yes**, what was the time of onset |  |  |
|  |  | □ < 1 month □ 1-2 months □ >2 -3 months □ >3-4 months □ >4-5 months  □ >5-6 months □ >6-7 months □ >7 -8 months □ >8-9 months □ >9 -10 months  □ >10-11 months □ >11 -12 months □ >12 months □ From the time of discharge  □ Not sure |  |  |
|  |  | **If yes**, what was the duration of symptoms |  |  |
|  |  | □ < 1 month □ 1-2 months □ >2 -3 months □ >3-4 months □ >4-5 months  □ >5-6 months □ >6-7 months □ >7 -8 months □ >8-9 months □ >9 -10 months  □ >10-11 months □ >11 -12 months □ >12 months □ Not sure |  |  |
| Chest pain | □ Yes □ No | **If yes**, what was the time of onset |  |  |
|  |  | □ < 1 month □ 1-2 months □ >2 -3 months □ >3-4 months □ >4-5 months  □ >5-6 months □ >6-7 months □ >7 -8 months □ >8-9 months □ >9 -10 months  □ >10-11 months □ >11 -12 months □ >12 months □ From the time of discharge  □ Not sure |  |  |
|  |  | **If yes**, what was the duration of symptoms |  |  |
|  |  | □ < 1 month □ 1-2 months □ >2 -3 months □ >3-4 months □ >4-5 months  □ >5-6 months □ >6-7 months □ >7 -8 months □ >8-9 months □ >9 -10 months  □ >10-11 months □ >11 -12 months □ >12 months □ Not sure |  |  |
| Persistent cough | □ Yes □ No | **If yes**, what was the time of onset |  |  |
|  |  | □ < 1 month □ 1-2 months □ >2 -3 months □ >3-4 months □ >4-5 months  □ >5-6 months □ >6-7 months □ >7 -8 months □ >8-9 months □ >9 -10 months  □ >10-11 months □ >11 -12 months □ >12 months □ From the time of discharge  □ Not sure |  |  |
|  |  | **If yes**, what was the duration of symptoms |  |  |
|  |  | □ < 1 month □ 1-2 months □ >2 -3 months □ >3-4 months □ >4-5 months  □ >5-6 months □ >6-7 months □ >7 -8 months □ >8-9 months □ >9 -10 months  □ >10-11 months □ >11 -12 months □ >12 months □ Not sure |  |  |
| *If yes, □ dry cough □ with phlegm* | | |  |  |
| **Musculoskeletal problems** | **Tick Yes or No** | **If yes**, what was the time of onset |  |  |
| Cannot fully move or control movement | □ Yes □ No | □ < 1 month □ 1-2 months □ >2 -3 months □ >3-4 months □ >4-5 months  □ >5-6 months □ >6-7 months □ >7 -8 months □ >8-9 months □ >9 -10 months  □ >10-11 months □ >11 -12 months □ >12 months □ From the time of discharge  □ Not sure |  |  |
|  |  | **If yes**, what was the duration of symptoms |  |  |
|  |  | □ < 1 month □ 1-2 months □ >2 -3 months □ >3-4 months □ >4-5 months  □ >5-6 months □ >6-7 months □ >7 -8 months □ >8-9 months □ >9 -10 months  □ >10-11 months □ >11 -12 months □ >12 months □ Not sure |  |  |
| Problems with balance | □ Yes □ No | **If yes**, what was the time of onset |  |  |
|  |  | □ < 1 month □ 1-2 months □ >2 -3 months □ >3-4 months □ >4-5 months  □ >5-6 months □ >6-7 months □ >7 -8 months □ >8-9 months □ >9 -10 months  □ >10-11 months □ >11 -12 months □ >12 months □ From the time of discharge  □ Not sure |  |  |
|  |  | **If yes**, what was the duration of symptoms |  |  |
|  |  | □ < 1 month □ 1-2 months □ >2 -3 months □ >3-4 months □ >4-5 months  □ >5-6 months □ >6-7 months □ >7 -8 months □ >8-9 months □ >9 -10 months  □ >10-11 months □ >11 -12 months □ >12 months □ Not sure |  |  |
| Persistent muscle pain | □ Yes □ No | **If yes**, what was the time of onset |  |  |
|  |  | □ < 1 month □ 1-2 months □ >2 -3 months □ >3-4 months □ >4-5 months  □ >5-6 months □ >6-7 months □ >7 -8 months □ >8-9 months □ >9 -10 months  □ >10-11 months □ >11 -12 months □ >12 months □ From the time of discharge  □ Not sure |  |  |
|  |  | **If yes**, what was the duration of symptoms |  |  |
|  |  | □ < 1 month □ 1-2 months □ >2 -3 months □ >3-4 months □ >4-5 months  □ >5-6 months □ >6-7 months □ >7 -8 months □ >8-9 months □ >9 -10 months  □ >10-11 months □ >11 -12 months □ >12 months □ Not sure |  |  |
| Joint pain or swelling | □ Yes □ No | **If yes**, what was the time of onset |  |  |
|  |  | □ < 1 month □ 1-2 months □ >2 -3 months □ >3-4 months □ >4-5 months  □ >5-6 months □ >6-7 months □ >7 -8 months □ >8-9 months □ >9 -10 months  □ >10-11 months □ >11 -12 months □ >12 months □ From the time of discharge  □ Not sure |  |  |
|  |  | **If yes**, what was the duration of symptoms |  |  |
|  |  | □ < 1 month □ 1-2 months □ >2 -3 months □ >3-4 months □ >4-5 months  □ >5-6 months □ >6-7 months □ >7 -8 months □ >8-9 months □ >9 -10 months  □ >10-11 months □ >11 -12 months □ >12 months □ Not sure |  |  |
| **Neurological problems** | **Tick Yes or No** | **If yes**, what was the time of onset |  |  |
| Headache | □ Yes □ No | □ < 1 month □ 1-2 months □ >2 -3 months □ >3-4 months □ >4-5 months  □ >5-6 months □ >6-7 months □ >7 -8 months □ >8-9 months □ >9 -10 months  □ >10-11 months □ >11 -12 months □ >12 months □ From the time of discharge  □ Not sure |  |  |
|  |  | **If yes**, what was the duration of symptoms |  |  |
|  |  | □ < 1 month □ 1-2 months □ >2 -3 months □ >3-4 months □ >4-5 months  □ >5-6 months □ >6-7 months □ >7 -8 months □ >8-9 months □ >9 -10 months  □ >10-11 months □ >11 -12 months □ >12 months □ Not sure |  |  |
| Dizziness/ light headedness | □ Yes □ No | **If yes**, what was the time of onset |  |  |
|  |  | □ < 1 month □ 1-2 months □ >2 -3 months □ >3-4 months □ >4-5 months  □ >5-6 months □ >6-7 months □ >7 -8 months □ >8-9 months □ >9 -10 months  □ >10-11 months □ >11 -12 months □ >12 months □ From the time of discharge  □ Not sure |  |  |
|  |  | **If yes**, what was the duration of symptoms |  |  |
|  |  | □ < 1 month □ 1-2 months □ >2 -3 months □ >3-4 months □ >4-5 months  □ >5-6 months □ >6-7 months □ >7 -8 months □ >8-9 months □ >9 -10 months  □ >10-11 months □ >11 -12 months □ >12 months □ Not sure |  |  |
| Fainting/ blackouts | □ Yes □ No | **If yes**, what was the time of onset |  |  |
|  |  | □ < 1 month □ 1-2 months □ >2 -3 months □ >3-4 months □ >4-5 months  □ >5-6 months □ >6-7 months □ >7 -8 months □ >8-9 months □ >9 -10 months  □ >10-11 months □ >11 -12 months □ >12 months □ From the time of discharge  □ Not sure |  |  |
|  |  | **If yes**, what was the duration of symptoms |  |  |
|  |  | □ < 1 month □ 1-2 months □ >2 -3 months □ >3-4 months □ >4-5 months  □ >5-6 months □ >6-7 months □ >7 -8 months □ >8-9 months □ >9 -10 months  □ >10-11 months □ >11 -12 months □ >12 months □ Not sure |  |  |
| Problems seeing/blurred vision | □ Yes □ No | **If yes**, what was the time of onset |  |  |
|  |  | □ < 1 month □ 1-2 months □ >2 -3 months □ >3-4 months □ >4-5 months  □ >5-6 months □ >6-7 months □ >7 -8 months □ >8-9 months □ >9 -10 months  □ >10-11 months □ >11 -12 months □ >12 months □ From the time of discharge  □ Not sure |  |  |
|  |  | **If yes**, what was the duration of symptoms |  |  |
|  |  | □ < 1 month □ 1-2 months □ >2 -3 months □ >3-4 months □ >4-5 months  □ >5-6 months □ >6-7 months □ >7 -8 months □ >8-9 months □ >9 -10 months  □ >10-11 months □ >11 -12 months □ >12 months □ Not sure |  |  |
| Disturbed smell | □ Yes □ No | **If yes**, what was the time of onset |  |  |
|  |  | □ < 1 month □ 1-2 months □ >2 -3 months □ >3-4 months □ >4-5 months  □ >5-6 months □ >6-7 months □ >7 -8 months □ >8-9 months □ >9 -10 months  □ >10-11 months □ >11 -12 months □ >12 months □ From the time of discharge  □ Not sure |  |  |
|  |  | **If yes**, what was the duration of symptoms |  |  |
|  |  | □ < 1 month □ 1-2 months □ >2 -3 months □ >3-4 months □ >4-5 months  □ >5-6 months □ >6-7 months □ >7 -8 months □ >8-9 months □ >9 -10 months  □ >10-11 months □ >11 -12 months □ >12 months □ Not sure |  |  |
| Loss of smell | □ Yes □ No | **If yes**, what was the time of onset |  |  |
|  |  | □ < 1 month □ 1-2 months □ >2 -3 months □ >3-4 months □ >4-5 months  □ >5-6 months □ >6-7 months □ >7 -8 months □ >8-9 months □ >9 -10 months  □ >10-11 months □ >11 -12 months □ >12 months □ From the time of discharge  □ Not sure |  |  |
|  |  | **If yes**, what was the duration of symptoms |  |  |
|  |  | □ < 1 month □ 1-2 months □ >2 -3 months □ >3-4 months □ >4-5 months  □ >5-6 months □ >6-7 months □ >7 -8 months □ >8-9 months □ >9 -10 months  □ >10-11 months □ >11 -12 months □ >12 months □ Not sure |  |  |
| Disturbed taste | □ Yes □ No | **If yes**, what was the time of onset |  |  |
|  |  | □ < 1 month □ 1-2 months □ >2 -3 months □ >3-4 months □ >4-5 months  □ >5-6 months □ >6-7 months □ >7 -8 months □ >8-9 months □ >9 -10 months  □ >10-11 months □ >11 -12 months □ >12 months □ From the time of discharge  □ Not sure |  |  |
|  |  | **If yes**, what was the duration of symptoms |  |  |
|  |  | □ < 1 month □ 1-2 months □ >2 -3 months □ >3-4 months □ >4-5 months  □ >5-6 months □ >6-7 months □ >7 -8 months □ >8-9 months □ >9 -10 months  □ >10-11 months □ >11 -12 months □ >12 months □ Not sure |  |  |
| Loss of taste | □ Yes □ No | **If yes**, what was the time of onset |  |  |
|  |  | □ < 1 month □ 1-2 months □ >2 -3 months □ >3-4 months □ >4-5 months  □ >5-6 months □ >6-7 months □ >7 -8 months □ >8-9 months □ >9 -10 months  □ >10-11 months □ >11 -12 months □ >12 months □ From the time of discharge  □ Not sure |  |  |
|  |  | **If yes**, what was the duration of symptoms |  |  |
|  |  | □ < 1 month □ 1-2 months □ >2 -3 months □ >3-4 months □ >4-5 months  □ >5-6 months □ >6-7 months □ >7 -8 months □ >8-9 months □ >9 -10 months  □ >10-11 months □ >11 -12 months □ >12 months □ Not sure |  |  |
| Tremor/shakiness | □ Yes □ No | **If yes**, what was the time of onset |  |  |
|  |  | □ < 1 month □ 1-2 months □ >2 -3 months □ >3-4 months □ >4-5 months  □ >5-6 months □ >6-7 months □ >7 -8 months □ >8-9 months □ >9 -10 months  □ >10-11 months □ >11 -12 months □ >12 months □ From the time of discharge  □ Not sure |  |  |
|  |  | **If yes**, what was the duration of symptoms |  |  |
|  |  | □ < 1 month □ 1-2 months □ >2 -3 months □ >3-4 months □ >4-5 months  □ >5-6 months □ >6-7 months □ >7 -8 months □ >8-9 months □ >9 -10 months  □ >10-11 months □ >11 -12 months □ >12 months □ Not sure |  |  |
| Tingling feeling/ “pins and needles“ | □ Yes □ No | **If yes**, what was the time of onset |  |  |
|  |  | □ < 1 month □ 1-2 months □ >2 -3 months □ >3-4 months □ >4-5 months  □ >5-6 months □ >6-7 months □ >7 -8 months □ >8-9 months □ >9 -10 months  □ >10-11 months □ >11 -12 months □ >12 months □ From the time of discharge  □ Not sure |  |  |
|  |  | **If yes**, what was the duration of symptoms |  |  |
|  |  | □ < 1 month □ 1-2 months □ >2 -3 months □ >3-4 months □ >4-5 months  □ >5-6 months □ >6-7 months □ >7 -8 months □ >8-9 months □ >9 -10 months  □ >10-11 months □ >11 -12 months □ >12 months □ Not sure |  |  |
| Seizures/fits | □ Yes □ No | **If yes**, what was the time of onset |  |  |
|  |  | □ < 1 month □ 1-2 months □ >2 -3 months □ >3-4 months □ >4-5 months  □ >5-6 months □ >6-7 months □ >7 -8 months □ >8-9 months □ >9 -10 months  □ >10-11 months □ >11 -12 months □ >12 months □ From the time of discharge  □ Not sure |  |  |
|  |  | **If yes**, what was the duration of symptoms |  |  |
|  |  | □ < 1 month □ 1-2 months □ >2 -3 months □ >3-4 months □ >4-5 months  □ >5-6 months □ >6-7 months □ >7 -8 months □ >8-9 months □ >9 -10 months  □ >10-11 months □ >11 -12 months □ >12 months □ Not sure |  |  |
| Confusion/lack of concentration | □ Yes □ No | **If yes**, what was the time of onset |  |  |
|  |  | □ < 1 month □ 1-2 months □ >2 -3 months □ >3-4 months □ >4-5 months  □ >5-6 months □ >6-7 months □ >7 -8 months □ >8-9 months □ >9 -10 months  □ >10-11 months □ >11 -12 months □ >12 months □ From the time of discharge  □ Not sure |  |  |
|  |  | **If yes**, what was the duration of symptoms |  |  |
|  |  | □ < 1 month □ 1-2 months □ >2 -3 months □ >3-4 months □ >4-5 months  □ >5-6 months □ >6-7 months □ >7 -8 months □ >8-9 months □ >9 -10 months  □ >10-11 months □ >11 -12 months □ >12 months □ Not sure |  |  |
| Problems speaking or communicating | □ Yes □ No | **If yes**, what was the time of onset |  |  |
|  |  | □ < 1 month □ 1-2 months □ >2 -3 months □ >3-4 months □ >4-5 months  □ >5-6 months □ >6-7 months □ >7 -8 months □ >8-9 months □ >9 -10 months  □ >10-11 months □ >11 -12 months □ >12 months □ From the time of discharge  □ Not sure |  |  |
|  |  | **If yes**, what was the duration of symptoms |  |  |
|  |  | □ < 1 month □ 1-2 months □ >2 -3 months □ >3-4 months □ >4-5 months  □ >5-6 months □ >6-7 months □ >7 -8 months □ >8-9 months □ >9 -10 months  □ >10-11 months □ >11 -12 months □ >12 months □ Not sure |  |  |
| Insomnia *(hard to fall asleep, hard to stay asleep)* | □ Yes □ No | **If yes**, what was the time of onset |  |  |
|  |  | □ < 1 month □ 1-2 months □ >2 -3 months □ >3-4 months □ >4-5 months  □ >5-6 months □ >6-7 months □ >7 -8 months □ >8-9 months □ >9 -10 months  □ >10-11 months □ >11 -12 months □ >12 months □ From the time of discharge  □ Not sure |  |  |
|  |  | **If yes**, what was the duration of symptoms |  |  |
|  |  | □ < 1 month □ 1-2 months □ >2 -3 months □ >3-4 months □ >4-5 months  □ >5-6 months □ >6-7 months □ >7 -8 months □ >8-9 months □ >9 -10 months  □ >10-11 months □ >11 -12 months □ >12 months □ Not sure |  |  |
| Hypersomnia *(excessive daytime sleepiness or prolonged nighttime sleep)* | □ Yes □ No | **If yes**, what was the time of onset |  |  |
|  |  | □ < 1 month □ 1-2 months □ >2 -3 months □ >3-4 months □ >4-5 months  □ >5-6 months □ >6-7 months □ >7 -8 months □ >8-9 months □ >9 -10 months  □ >10-11 months □ >11 -12 months □ >12 months □ From the time of discharge  □ Not sure |  |  |
|  |  | **If yes**, what was the duration of symptoms |  |  |
|  |  | □ < 1 month □ 1-2 months □ >2 -3 months □ >3-4 months □ >4-5 months  □ >5-6 months □ >6-7 months □ >7 -8 months □ >8-9 months □ >9 -10 months  □ >10-11 months □ >11 -12 months □ >12 months □ Not sure |  |  |
| **Fatigue** | □ Yes □ No | **If yes**, what was the time of onset |  |  |
|  |  | □ < 1 month □ 1-2 months □ >2 -3 months □ >3-4 months □ >4-5 months  □ >5-6 months □ >6-7 months □ >7 -8 months □ >8-9 months □ >9 -10 months  □ >10-11 months □ >11 -12 months □ >12 months □ From the time of discharge  □ Not sure |  |  |
|  |  | **If yes**, what was the duration of symptoms |  |  |
|  |  | □ < 1 month □ 1-2 months □ >2 -3 months □ >3-4 months □ >4-5 months  □ >5-6 months □ >6-7 months □ >7 -8 months □ >8-9 months □ >9 -10 months  □ >10-11 months □ >11 -12 months □ >12 months □ Not sure |  |  |
| **Gastrointestinal problems** | **Tick Yes or No** | **If yes**, what was the time of onset |  |  |
| Weight loss | □ Yes □ No | □ < 1 month □ 1-2 months □ >2 -3 months □ >3-4 months □ >4-5 months  □ >5-6 months □ >6-7 months □ >7 -8 months □ >8-9 months □ >9 -10 months  □ >10-11 months □ >11 -12 months □ >12 months □ From the time of discharge  □ Not sure |  |  |
|  |  | **If yes**, what was the duration of symptoms |  |  |
|  |  | □ < 1 month □ 1-2 months □ >2 -3 months □ >3-4 months □ >4-5 months  □ >5-6 months □ >6-7 months □ >7 -8 months □ >8-9 months □ >9 -10 months  □ >10-11 months □ >11 -12 months □ >12 months □ Not sure |  |  |
| Problems swallowing or chewing | □ Yes □ No | **If yes**, what was the time of onset |  |  |
|  |  | □ < 1 month □ 1-2 months □ >2 -3 months □ >3-4 months □ >4-5 months  □ >5-6 months □ >6-7 months □ >7 -8 months □ >8-9 months □ >9 -10 months  □ >10-11 months □ >11 -12 months □ >12 months □ From the time of discharge  □ Not sure |  |  |
|  |  | **If yes**, what was the duration of symptoms |  |  |
|  |  | □ < 1 month □ 1-2 months □ >2 -3 months □ >3-4 months □ >4-5 months  □ >5-6 months □ >6-7 months □ >7 -8 months □ >8-9 months □ >9 -10 months  □ >10-11 months □ >11 -12 months □ >12 months □ Not sure |  |  |
| Poor appetite | □ Yes □ No | **If yes**, what was the time of onset |  |  |
|  |  | □ < 1 month □ 1-2 months □ >2 -3 months □ >3-4 months □ >4-5 months  □ >5-6 months □ >6-7 months □ >7 -8 months □ >8-9 months □ >9 -10 months  □ >10-11 months □ >11 -12 months □ >12 months □ From the time of discharge  □ Not sure |  |  |
|  |  | **If yes**, what was the duration of symptoms |  |  |
|  |  | □ < 1 month □ 1-2 months □ >2 -3 months □ >3-4 months □ >4-5 months  □ >5-6 months □ >6-7 months □ >7 -8 months □ >8-9 months □ >9 -10 months  □ >10-11 months □ >11 -12 months □ >12 months □ Not sure |  |  |
| Diarrhea | □ Yes □ No | **If yes**, what was the time of onset |  |  |
|  |  | □ < 1 month □ 1-2 months □ >2 -3 months □ >3-4 months □ >4-5 months  □ >5-6 months □ >6-7 months □ >7 -8 months □ >8-9 months □ >9 -10 months  □ >10-11 months □ >11 -12 months □ >12 months □ From the time of discharge  □ Not sure |  |  |
|  |  | **If yes**, what was the duration of symptoms |  |  |
|  |  | □ < 1 month □ 1-2 months □ >2 -3 months □ >3-4 months □ >4-5 months  □ >5-6 months □ >6-7 months □ >7 -8 months □ >8-9 months □ >9 -10 months  □ >10-11 months □ >11 -12 months □ >12 months □ Not sure |  |  |
| Stomach/ abdominal pain | □ Yes □ No | **If yes**, what was the time of onset |  |  |
|  |  | □ < 1 month □ 1-2 months □ >2 -3 months □ >3-4 months □ >4-5 months  □ >5-6 months □ >6-7 months □ >7 -8 months □ >8-9 months □ >9 -10 months  □ >10-11 months □ >11 -12 months □ >12 months □ From the time of discharge  □ Not sure |  |  |
|  |  | **If yes**, what was the duration of symptoms |  |  |
|  |  | □ < 1 month □ 1-2 months □ >2 -3 months □ >3-4 months □ >4-5 months  □ >5-6 months □ >6-7 months □ >7 -8 months □ >8-9 months □ >9 -10 months  □ >10-11 months □ >11 -12 months □ >12 months □ Not sure |  |  |
| Feeling nauseous | □ Yes □ No | **If yes**, what was the time of onset |  |  |
|  |  | □ < 1 month □ 1-2 months □ >2 -3 months □ >3-4 months □ >4-5 months  □ >5-6 months □ >6-7 months □ >7 -8 months □ >8-9 months □ >9 -10 months  □ >10-11 months □ >11 -12 months □ >12 months □ From the time of discharge  □ Not sure |  |  |
|  |  | **If yes**, what was the duration of symptoms |  |  |
|  |  | □ < 1 month □ 1-2 months □ >2 -3 months □ >3-4 months □ >4-5 months  □ >5-6 months □ >6-7 months □ >7 -8 months □ >8-9 months □ >9 -10 months  □ >10-11 months □ >11 -12 months □ >12 months □ Not sure |  |  |
| Vomiting | □ Yes □ No | **If yes**, what was the time of onset |  |  |
|  |  | □ < 1 month □ 1-2 months □ >2 -3 months □ >3-4 months □ >4-5 months  □ >5-6 months □ >6-7 months □ >7 -8 months □ >8-9 months □ >9 -10 months  □ >10-11 months □ >11 -12 months □ >12 months □ From the time of discharge  □ Not sure |  |  |
|  |  | **If yes**, what was the duration of symptoms |  |  |
|  |  | □ < 1 month □ 1-2 months □ >2 -3 months □ >3-4 months □ >4-5 months  □ >5-6 months □ >6-7 months □ >7 -8 months □ >8-9 months □ >9 -10 months  □ >10-11 months □ >11 -12 months □ >12 months □ Not sure |  |  |
| Constipation | □ Yes □ No | **If yes**, what was the time of onset |  |  |
|  |  | □ < 1 month □ 1-2 months □ >2 -3 months □ >3-4 months □ >4-5 months  □ >5-6 months □ >6-7 months □ >7 -8 months □ >8-9 months □ >9 -10 months  □ >10-11 months □ >11 -12 months □ >12 months □ From the time of discharge  □ Not sure |  |  |
|  |  | **If yes**, what was the duration of symptoms |  |  |
|  |  | □ < 1 month □ 1-2 months □ >2 -3 months □ >3-4 months □ >4-5 months  □ >5-6 months □ >6-7 months □ >7 -8 months □ >8-9 months □ >9 -10 months  □ >10-11 months □ >11 -12 months □ >12 months □ Not sure |  |  |
| **Cardiovascular problems** | **Tick Yes or No** | **If yes**, what was the time of onset |  |  |
| Palpitations (heart racing) | □ Yes □ No | □ < 1 month □ 1-2 months □ >2 -3 months □ >3-4 months □ >4-5 months  □ >5-6 months □ >6-7 months □ >7 -8 months □ >8-9 months □ >9 -10 months  □ >10-11 months □ >11 -12 months □ >12 months □ From the time of discharge  □ Not sure |  |  |
|  |  | **If yes**, what was the duration of symptoms |  |  |
|  |  | □ < 1 month □ 1-2 months □ >2 -3 months □ >3-4 months □ >4-5 months  □ >5-6 months □ >6-7 months □ >7 -8 months □ >8-9 months □ >9 -10 months  □ >10-11 months □ >11 -12 months □ >12 months □ Not sure |  |  |
| Variations in heart rate (tachycardia or bradycardia) | □ Yes □ No | **If yes**, what was the time of onset |  |  |
|  |  | □ < 1 month □ 1-2 months □ >2 -3 months □ >3-4 months □ >4-5 months  □ >5-6 months □ >6-7 months □ >7 -8 months □ >8-9 months □ >9 -10 months  □ >10-11 months □ >11 -12 months □ >12 months □ From the time of discharge  □ Not sure |  |  |
|  |  | **If yes**, what was the duration of symptoms |  |  |
|  |  | □ < 1 month □ 1-2 months □ >2 -3 months □ >3-4 months □ >4-5 months  □ >5-6 months □ >6-7 months □ >7 -8 months □ >8-9 months □ >9 -10 months  □ >10-11 months □ >11 -12 months □ >12 months □ Not sure |  |  |
| Bleeding | □ Yes □ No | **If yes**, what was the time of onset |  |  |
|  |  | □ < 1 month □ 1-2 months □ >2 -3 months □ >3-4 months □ >4-5 months  □ >5-6 months □ >6-7 months □ >7 -8 months □ >8-9 months □ >9 -10 months  □ >10-11 months □ >11 -12 months □ >12 months □ From the time of discharge  □ Not sure |  |  |
|  |  | **If yes**, what was the duration of symptoms |  |  |
|  |  | □ < 1 month □ 1-2 months □ >2 -3 months □ >3-4 months □ >4-5 months  □ >5-6 months □ >6-7 months □ >7 -8 months □ >8-9 months □ >9 -10 months  □ >10-11 months □ >11 -12 months □ >12 months □ Not sure |  |  |
| *If yes, specify bleeding site:* |  |  |  |  |
| **Genitourinary problems** | **Tick Yes or No** | **If yes**, what was the time of onset |  | **If yes**, what is the duration of symptoms |
| Urination problems | □ Yes □ No | □ < 1 month □ 1-2 months □ >2 -3 months □ >3-4 months □ >4-5 months  □ >5-6 months □ >6-7 months □ >7 -8 months □ >8-9 months □ >9 -10 months  □ >10-11 months □ >11 -12 months □ >12 months □ From the time of discharge  □ Not sure |  |  |
|  |  | **If yes**, what was the duration of symptoms |  |  |
|  |  | □ < 1 month □ 1-2 months □ >2 -3 months □ >3-4 months □ >4-5 months  □ >5-6 months □ >6-7 months □ >7 -8 months □ >8-9 months □ >9 -10 months  □ >10-11 months □ >11 -12 months □ >12 months □ Not sure |  |  |
| Changes in menstruation, (if regular before Covid-19 illness) | □ Yes □ No  □ Not applicable | **If yes**, what was the time of onset |  |  |
|  |  | □ < 1 month □ 1-2 months □ >2 -3 months □ >3-4 months □ >4-5 months  □ >5-6 months □ >6-7 months □ >7 -8 months □ >8-9 months □ >9 -10 months  □ >10-11 months □ >11 -12 months □ >12 months □ From the time of discharge  □ Not sure |  |  |
|  |  | **If yes**, what was the duration of symptoms |  |  |
|  |  | □ < 1 month □ 1-2 months □ >2 -3 months □ >3-4 months □ >4-5 months  □ >5-6 months □ >6-7 months □ >7 -8 months □ >8-9 months □ >9 -10 months  □ >10-11 months □ >11 -12 months □ >12 months □ Not sure |  |  |
| **Other problems** | **Tick Yes or No** | **If yes**, what was the time of onset |  |  |
| Bilateral conjunctivitis  *If yes, □ purulent □ non-purulent* | □ Yes □ No | □ < 1 month □ 1-2 months □ >2 -3 months □ >3-4 months □ >4-5 months  □ >5-6 months □ >6-7 months □ >7 -8 months □ >8-9 months □ >9 -10 months  □ >10-11 months □ >11 -12 months □ >12 months □ From the time of discharge  □ Not sure |  |  |
|  |  | **If yes**, what was the duration of symptoms |  |  |
|  |  | □ < 1 month □ 1-2 months □ >2 -3 months □ >3-4 months □ >4-5 months  □ >5-6 months □ >6-7 months □ >7 -8 months □ >8-9 months □ >9 -10 months  □ >10-11 months □ >11 -12 months □ >12 months □ Not sure |  |  |
| Lumps or rashes (purple/pink) on toes | □ Yes □ No | **If yes**, what was the time of onset |  |  |
|  |  | □ < 1 month □ 1-2 months □ >2 -3 months □ >3-4 months □ >4-5 months  □ >5-6 months □ >6-7 months □ >7 -8 months □ >8-9 months □ >9 -10 months  □ >10-11 months □ >11 -12 months □ >12 months □ From the time of discharge  □ Not sure |  |  |
|  |  | **If yes**, what was the duration of symptoms |  |  |
|  |  | □ < 1 month □ 1-2 months □ >2 -3 months □ >3-4 months □ >4-5 months  □ >5-6 months □ >6-7 months □ >7 -8 months □ >8-9 months □ >9 -10 months  □ >10-11 months □ >11 -12 months □ >12 months □ Not sure |  |  |
| Skin rash | □ Yes □ No | **If yes**, what was the time of onset |  |  |
|  |  | □ < 1 month □ 1-2 months □ >2 -3 months □ >3-4 months □ >4-5 months  □ >5-6 months □ >6-7 months □ >7 -8 months □ >8-9 months □ >9 -10 months  □ >10-11 months □ >11 -12 months □ >12 months □ From the time of discharge  □ Not sure |  |  |
|  |  | **If yes**, what was the duration of symptoms |  |  |
|  |  | □ < 1 month □ 1-2 months □ >2 -3 months □ >3-4 months □ >4-5 months  □ >5-6 months □ >6-7 months □ >7 -8 months □ >8-9 months □ >9 -10 months  □ >10-11 months □ >11 -12 months □ >12 months □ Not sure |  |  |
| Skin rash *If yes, tick all body areas that apply:* | □ Yes □ No | □ *Face* |  |  |
|  |  | □ *Trunk (stomach or back)* |  |  |
|  |  | □ *Arms* |  |  |
|  |  | □ *Legs* |  |  |
|  |  | □ *Buttocks* |  |  |
|  |  | □ *Toes* |  |  |
|  |  | □ *Fingers* |  |  |
|  |  | □ *Accompanied by itch* |  |  |
| Hair loss | □ Yes □ No | **If yes**, what was the time of onset |  |  |
|  |  | □ < 1 month □ 1-2 months □ >2 -3 months □ >3-4 months □ >4-5 months  □ >5-6 months □ >6-7 months □ >7 -8 months □ >8-9 months □ >9 -10 months  □ >10-11 months □ >11 -12 months □ >12 months □ From the time of discharge  □ Not sure |  |  |
|  |  | **If yes**, what was the duration of symptoms |  |  |
|  |  | □ < 1 month □ 1-2 months □ >2 -3 months □ >3-4 months □ >4-5 months  □ >5-6 months □ >6-7 months □ >7 -8 months □ >8-9 months □ >9 -10 months  □ >10-11 months □ >11 -12 months □ >12 months □ Not sure |  |  |
| Hyperhidrosis | □ Yes □ No | **If yes**, what was the time of onset |  |  |
|  |  | □ < 1 month □ 1-2 months □ >2 -3 months □ >3-4 months □ >4-5 months  □ >5-6 months □ >6-7 months □ >7 -8 months □ >8-9 months □ >9 -10 months  □ >10-11 months □ >11 -12 months □ >12 months □ From the time of discharge  □ Not sure |  |  |
|  |  | **If yes**, what was the duration of symptoms |  |  |
|  |  | □ < 1 month □ 1-2 months □ >2 -3 months □ >3-4 months □ >4-5 months  □ >5-6 months □ >6-7 months □ >7 -8 months □ >8-9 months □ >9 -10 months  □ >10-11 months □ >11 -12 months □ >12 months □ Not sure |  |  |
| **Other New Symptoms, if yes, specify all with their onset and duration:** | | **If yes**, what was the time of onset |  |  |
|  | | □ < 1 month □ 1-2 months □ >2 -3 months □ >3-4 months □ >4-5 months  □ >5-6 months □ >6-7 months □ >7 -8 months □ >8-9 months □ >9 -10 months  □ >10-11 months □ >11 -12 months □ >12 months □ From the time of discharge  □ Not sure |  |  |
|  |  | **If yes**, what was the duration of symptoms |  |  |
|  |  | □ < 1 month □ 1-2 months □ >2 -3 months □ >3-4 months □ >4-5 months  □ >5-6 months □ >6-7 months □ >7 -8 months □ >8-9 months □ >9 -10 months  □ >10-11 months □ >11 -12 months □ >12 months □ Not sure |  |  |

| **7. Your child’s overall health status** | | |
| --- | --- | --- |
| We would like to know how good or bad your child’s health was  **BEFORE Covid-19** and how it is **TODAY**  This scale is numbered from 0 to 100%  with **100% meaning the best health** you can imagine  **0% means the worst health** you can imagine.  Please indicate on the scale and **write the number in the box below each scale** to indicate how good or bad your child’s health was **BEFORE Covid-19** and how it is **TODAY**. | Best health | Best health |
|  | 100; 95; 90; 85; 80; 75; 70; 65; 60; 55; 50; 45; 40; 35; 30; 25; 20; 15; 10; 5; 0 **Before Covid-19** | 100; 95; 90; 85; 80; 75; 70; 65; 60; 55; 50; 45; 40; 35; 30; 25; 20; 15; 10; 5; 0 **Today** |

**8. Vaccinations**

**Has your child been vaccinated in accordance with the national vaccination schedule?**

□ Yes, vaccinated up to date □ Yes, but some vaccines were missed □ No, I avoid vaccination for my child

**Please provide an approximate date of your child’s latest vaccination?** [_D_][_D_]/[_M_][_M_]/[_2_][_0_][_Y_][_Y_]

Please, specify what was the vaccine: __________________□ I do not remember

**I trust information I receive about vaccines?**
 □ Not at all □ Only a little □ Undecided □ Quite a lot □ A great deal

**How confident are you in any of the childhood vaccines safety?**

□ Not at all □ Only a little □ Undecided □ Quite a lot □ A great deal

**Has your child been vaccinated against Covid-19?** □ Yes □ No □ Not sure

If yes, how many times have they had the Covid-19 vaccine? [_Number_]

Estimated date of the last vaccine dose received: [_D_][_D_]/[_M_][_M_]/[_2_][_0_][_2_][_Y_]

Which type of Covid-19 vaccine did they receive: AstraZeneca Pfizer-BioNTech

Imperial Janssens Moderna’s Sinopharm Sputnik V Other (name): ________ Not sure

**If no**, **would you like to vaccinate your child against Covid-19 in the future?** □ Yes □ No □ Not sure

**I trust information I receive about Covid-19 vaccination?**

□ Not at all □ Only a little □ Undecided □ Quite a lot □ A great deal

**How confident are you in the safety of Covid-19 vaccinations?**

□ Not at all □ Only a little □ Undecided □ Quite a lot □ A great deal

**What is your opinion of Russian-made vaccines against Covid-19?**

□ Negative □ Neutral □ Positive □ Not sure

**What is your opinion of vaccines against Covid-19 produced abroad?**

□ Negative □ Neutral □ Positive □ Not sure

| 9. **Some questions about you** |
| --- |

| During your child's illness, have you often been in a bad mood, depressed or feeling hopeless? | □ Yes □ No |
| --- | --- |
| During your child's illness, did you often feel that everything was difficult, and you did not want to do anything? | □ Yes □ No |
| During your child's illness, did you often feel persistent fatigue for no reason? | □ Yes □ No |
| Did your child's illness often make you feel nervous, anxious or extremely stressed? | □ Yes □ No |
| Did your child's illness often leave you unable to calm down or have you often been unable to calm or control your worries? | □ Yes □ No |
| Due to your child's illness, have you often experienced fear, as if something terrible were about to happen? | □ Yes □ No |
| Due to your child's illness, have you had to face aggressive or prejudiced attitudes from others? | □ Yes □ No |
| Did you receive enough help and support during your child's illness? | □ Yes □ No |

**10. Please let us know of any further comments about the child’s illness, the pandemic, lockdown and/or any sequelae.**

**11. End of survey**

**Thank you for your time!**

### **Table S1**. Categorisation of long-lasting symptoms at follow-up.

| Symptom category | Long-lasting symptoms included |
| --- | --- |
| Musculoskeletal | joint pain or swelling OR persistent muscle pain |
| Cardiovascular | variations in heart rate OR palpitations |
| Respiratory | difficulty breathing/chest tightness OR pain on breathing OR persistent cough |
| Neurological | cannot fully move or control movement OR problems with balance OR confusion/lack of concentration OR problems speaking or communicating OR seizures/fits OR tingling feeling/ ‘pins and needles’ OR tremor/shakiness OR dizziness/light headedness OR fainting/ blackouts |
| Dermatological | skin rash OR lumps or rashes (purple/pink) on toes OR  hair loss |
| Gastrointestinal | constipation OR diarrhea OR feeling nauseous OR stomach/ abdominal pain OR vomiting |
| Sensory | disturbed smell OR disturbed taste OR loss of smell OR  loss of taste |
| Sleep | hypersomnia OR insomnia |
| Fatigue | fatigue |

### **Table S2**. Symptoms at the time of hospital admission.

| Characteristics | Results |
| --- | --- |
| History of fever | 427/511 (83.6%) |
| Cough | 284/510 (55.7%) |
| Fatigue | 197/506 (38.9%) |
| Rhinorrhoea | 278/512 (54.3%) |
| Shortness of breath | 77/513 (15%) |
| Disturbed smell or loss of smell | 64/456 (14%) |
| Sore throat | 67/487 (13.8%) |
| Lymphadenopathy | 52/512 (10.2%) |
| Headache | 40/465 (8.6%) |
| Diarrhoea | 43/511 (8.4%) |
| Skin rash | 41/512 (8%) |
| Wheezing | 39/512 (7.6%) |
| Vomiting / Nausea | 32/512 (6.2%) |
| Chest pain | 28/464 (6%) |
| Abdominal pain | 27/489 (5.5%) |
| Disturbed taste or Loss of taste | 16/456 (3.5%) |
| Muscle aches | 14/463 (3%) |
| Conjunctivitis | 10/512 (2%) |
| Joint pain | 5/461 (1.1%) |
| Ear pain | 3/463 (0.6%) |
| Seizures | 3/512 (0.6%) |
| Bleeding | 3/512 (0.6%) |
| Lower chest wall indrawing | 3/512 (0.6%) |
| Confusion | 2/511 (0.4%) |

The differing denominators used indicate missing data.

**Table S3**. Most commonly used treatments during the hospital stay.

| Characteristics | Total |
| --- | --- |
| Antiviral or COVID-19 targeted agent | 394/512 (77.0%) |
| Antibiotics | 380/513 (74.1%) |
| Mucolytics | 188/513 (36.7%) |
| Arbidol | 133/512 (26%) |
| Antifungal agent | 25/513 (4.9%) |
| Corticosteroid | 20/513 (3.9%) |
| Heparin | 17/513 (3.3%) |

The differing denominators used indicate missing data.

**Table S4**. Symptoms reported at the time of the follow-up interview and symptom duration (in months).

| Current symptom | Total number of patients with the symptom | Total number of patients with the long-lasting symptom | < 1 | 1-2 | > 2-3 | > 3-4 | > 4-5 | > 5-6 | > 6-7 | > 7-8 | > 8-9 | > 9-10 | From the time of discharge |
| --- | --- | --- | --- | --- | --- | --- | --- | --- | --- | --- | --- | --- | --- |
| Fatigue | 63/498 (12.65%) | 53/496 (10.69%) | 5/496 (1.01%) | 1/496 (0.2%) | 0/496 (0%) | 1/496 (0.2%) | 1/496 (0.2%) | 4/496 (0.81%) | 1/496 (0.2%) | 2/496 (0.4%) | 1/496 (0.2%) | 0/496 (0%) | 45/496 (9.07%) |
| Nasal congestion/  rhinorrhea | 43/505(8.51%) | 10/505 (1.98%) | 29/505 (5.74 %) | 2/505 (0.4%) | 1/505 (0.2 %) | 1/505 (0.2%) | 0/505 (0%) | 0/505 (0%) | 0/505 (0%) | 0/505 (0%) | 0/505 (0%) | 0/505 (0%) | 10/505 (1.98 %) |
| Insomnia | 32/501 (6.39%) | 26/501 (5.19%) | 2/501 (0.4%) | 2/501 (0.4%) | 1/501 (0.2%) | 0/501 (0%) | 1/501 (0.2%) | 2/501 (0.4%) | 1/501 (0.2%) | 1/501 (0.2%) | 0/501 (0%) | 0/501 (0%) | 22/501 (4.39%) |
| Disturbed smell | 26/468 (5.56%) | 22/467 (4.71%) | 0/467 (0%) | 2/467 (0.43%) | 1/467 (0.21%) | 0/467 (0%) | 0/467 (0%) | 3/467 (0.64%) | 0/467 (0%) | 2/467 (0.43%) | 0/467 (0%) | 0/467 (0%) | 17/467 (3.64%) |
| Headache | 24/488(4.92%) | 17/486 (3.5%) | 4/486 (0.82%) | 0/486 (0%) | 1/486 (0.21%) | 0/486 (0%) | 0/486 (0%) | 1/486 (0.21%) | 0/486 (0%) | 0/486 (0%) | 0/486 (0%) | 0/486 (0%) | 16/486 (3.29%) |
| Disturbed taste | 18/468 (3.85%) | 16/468 (3.42%) | 0/468 (0%) | 1/468 (0.21%) | 1/468 (0.21%) | 0/468 (0%) | 0/468 (0%) | 2/468 (0.43%) | 0/468 (0%) | 2/468 (0.43%) | 0/468 (0%) | 0/468 (0%) | 12/468 (2.56%) |
| Hyperhidrosis | 17/502(3.39%) | 13/502 (2.59%) | 1/502 (0.2%) | 2/502 (0.4%) | 0/502 (0%) | 1/502 (0.2%) | 0/502 (0%) | 0/502 (0%) | 0/502 (0%) | 1/502 (0.2%) | 0/502 (0%) | 0/502 (0%) | 12/502 (2.39%) |
| Persistent cough | 17/503 (3.38%) | 5/503 (0.99%) | 9/503 (1.79%) | 3/503 (0.6%) | 0/503 (0%) | 0/503 (0%) | 0/503 (0%) | 0/503 (0%) | 0/503 (0%) | 0/503 (0%) | 0/503 (0%) | 0/503 (0%) | 5/503 (0.99%) |
| Hypersomnia | 16/501 (3.19%) | 15/501 (2.99%) | 0/501 (0%) | 0/501 (0%) | 0/501 (0%) | 1/501 (0.2%) | 0/501 (0%) | 1/501 (0.2%) | 1/501 (0.2%) | 1/501 (0.2%) | 1/501 (0.2%) | 0/501 (0%) | 11/501 (2.2%) |
| Poor appetite | 15/500 (3%) | 12/500 (2.4%) | 2/500 (0.4%) | 0/500 (0%) | 1/500 (0.2%) | 0/500 (0%) | 0/500 (0%) | 1/500 (0.2%) | 0/500 (0%) | 1/500 (0.2%) | 1/500 (0.2%) | 0/500 (0%) | 9/500 (1.8%) |
| Skin rash | 13/497 (2.62%) | 8/497 (1.61%) | 3/497 (0.6%) | 0/497 (0%) | 1/497 (0.2%) | 0/497 (0%) | 1/497 (0.2%) | 2/497 (0.4%) | 0/497 (0%) | 0/497 (0%) | 0/497 (0%) | 0/497 (0%) | 6/497 (1.21%) |
| Diarrhea | 13/499 (2.61%) | 10/499 (2%) | 1/499 (0.2%) | 0/499 (0%) | 1/499 (0.2%) | 0/499 (0%) | 1/499 (0.2%) | 0/499 (0%) | 0/499 (0%) | 1/499 (0.2%) | 0/499 (0%) | 0/499 (0%) | 9/499 (1.8%) |
| Stomach/ abdominal pain | 13/499 (2.61%) | 10/499 (2%) | 1/499 (0.2%) | 1/499 (0.2%) | 1/499 (0.2%) | 0/499 (0%) | 0/499 (0%) | 0/499 (0%) | 0/499 (0%) | 1/499 (0.2%) | 0/499 (0%) | 0/499 (0%) | 9/499 (1.8%) |
| Problems seeing/  blurred vision | 12/480 (2.5%) | 10/479 (2.09%) | 0/479 (0%) | 0/479 (0%) | 1/479 (0.21%) | 0/479 (0%) | 0/479 (0%) | 1/479 (0.21%) | 1/479 (0.21%) | 0/479 (0%) | 0/479 (0%) | 0/479 (0%) | 8/479 (1.67%) |
| Hair loss | 12/501 (2.4%) | 9/501 (1.8%) | 0/501 (0%) | 1/501 (0.2%) | 2/501 (0.4%) | 0/501 (0%) | 0/501 (0%) | 1/501 (0.2%) | 1/501 (0.2%) | 1/501 (0.2%) | 0/501 (0%) | 0/501 (0%) | 6/501 (1.2%) |
| Dizziness/ light headedness | 10/486 (2.06%) | 5/484 (1.03%) | 2/484 (0.41%) | 1/484 (0.21%) | 0/484 (0%) | 0/484 (0%) | 0/484 (0%) | 0/484 (0%) | 0/484 (0%) | 0/484 (0%) | 0/484 (0%) | 0/484 (0%) | 5/484 (1.03%) |
| Joint pain or swelling | 10/493 (2.03%) | 6/492 (1.22%) | 1/492 (0.2%) | 2/492 (0.41%) | 0/492 (0%) | 0/492 (0%) | 0/492 (0%) | 0/492 (0%) | 0/492 (0%) | 1/492 (0.2%) | 0/492 (0%) | 0/492 (0%) | 5/492 (1.02%) |
| Variations in heart rate | 10/494 (2.02%) | 6/493 (1.22%) | 0/493 (0%) | 0/493 (0%) | 1/493 (0.2%) | 0/493 (0%) | 0/493 (0%) | 0/493 (0%) | 0/493 (0%) | 1/493 (0.2%) | 0/493 (0%) | 0/493 (0%) | 5/493 (1.01%) |
| Constipation | 9/500 (1.8%) | 8/500 (1.6%) | 0/500 (0%) | 0/500 (0%) | 0/500 (0%) | 1/500 (0.2%) | 0/500 (0%) | 0/500 (0%) | 0/500 (0%) | 0/500 (0%) | 0/500 (0%) | 0/500 (0%) | 8/500 (1.6%) |
| Loss of smell | 8/468 (1.71%) | 7/468 (1.5%) | 0/468 (0%) | 1/468 (0.21%) | 0/468 (0%) | 0/468 (0%) | 0/468 (0%) | 0/468 (0%) | 0/468 (0%) | 0/468 (0%) | 0/468 (0%) | 0/468 (0%) | 7/468 (1.5%) |
| Difficulty breathing /chest tightness | 8/503 (1.59%) | 7/503 (1.39%) | 1/503 (0.2 %) | 0/503 (0%) | 0/503 (0%) | 0/503 (0%) | 0/503 (0%) | 1/503 (0.2 %) | 0/503 (0%) | 0/503 (0%) | 0/503 (0%) | 0/503 (0%) | 6/503 (1.19 %) |
| Palpitations | 7/472(1.48%) | 5/471 (1.06%) | 0/471 (0%) | 0/471 (0%) | 1/471 (0.21%) | 0/471 (0%) | 0/471 (0%) | 0/471 (0%) | 0/471 (0%) | 1/471 (0.21%) | 0/471 (0%) | 0/471 (0%) | 4/471 (0.85%) |
| Feeling nauseous | 7/500 (1.4%) | 6/500 (1.2%) | 1/500 (0.2%) | 0/500 (0%) | 0/500 (0%) | 0/500 (0%) | 0/500 (0%) | 0/500 (0%) | 1/500 (0.2%) | 1/500 (0.2%) | 0/500 (0%) | 0/500 (0%) | 4/500 (0.8%) |
| Chest pain | 6/487(1.23%) | 3/487 (0.62%) | 2/487 (0.41 %) | 0/487 (0%) | 1/487 (0.21 %) | 0/487 (0%) | 0/487 (0%) | 0/487 (0%) | 0/487 (0%) | 0/487 (0%) | 0/487 (0%) | 0/487 (0%) | 3/487 (0.62 %) |
| Persistent muscle pain | 6/491(1.22%) | 4/490 (0.82%) | 1/490 (0.2%) | 0/490 (0%) | 0/490 (0%) | 0/490 (0%) | 0/490 (0%) | 0/490 (0%) | 0/490 (0%) | 1/490 (0.2%) | 0/490 (0%) | 0/490 (0%) | 3/490 (0.61%) |
| Problems with balance | 6/496(1.21%) | 2/494 (0.4%) | 1/494 (0.2%) | 1/494 (0.2%) | 0/494 (0%) | 0/494 (0%) | 0/494 (0%) | 0/494 (0%) | 0/494 (0%) | 0/494 (0%) | 0/494 (0%) | 0/494 (0%) | 2/494 (0.4%) |
| Urination problems | 4/496 (0.81%) | 3/496 (0.6%) | 1/496 (0.2%) | 0/496 (0%) | 0/496 (0%) | 0/496 (0%) | 0/496 (0%) | 0/496 (0%) | 0/496 (0%) | 0/496 (0%) | 0/496 (0%) | 1/496 (0.2%) | 2/496 (0.4%) |
| Vomiting | 4/500 (0.8%) | 4/500 (0.8%) | 0/500 (0%) | 0/500 (0%) | 0/500 (0%) | 0/500 (0%) | 0/500 (0%) | 0/500 (0%) | 0/500 (0%) | 0/500 (0%) | 1/500 (0.2%) | 0/500 (0%) | 3/500 (0.6%) |
| Confusion/ lack of concentration | 3/486 (0.62%) | 2/486 (0.41%) | 0/486 (0%) | 0/486 (0%) | 0/486 (0%) | 0/486 (0%) | 1/486 (0.21%) | 1/486 (0.21%) | 0/486 (0%) | 0/486 (0%) | 0/486 (0%) | 0/486 (0%) | 1/486 (0.21%) |
| Pain on breathing | 3/488(0.61%) | 2/488 (0.41%) | 0/488 (0%) | 0/488 (0%) | 1/488 (0.2 %) | 0/488 (0%) | 0/488 (0%) | 0/488 (0%) | 0/488 (0%) | 0/488 (0%) | 0/488 (0%) | 0/488 (0%) | 2/488 (0.41%) |
| Cannot fully move or control movement | 3/499(0.6%) | 2/499 (0.4%) | 0/499 (0%) | 1/499 (0.2%) | 0/499 (0%) | 0/499 (0%) | 0/499 (0%) | 0/499 (0%) | 0/499 (0%) | 0/499 (0%) | 0/499 (0%) | 0/499 (0%) | 2/499 (0.4%) |
| Tremor/  shakiness | 3/500 (0.6%) | 3/500 (0.6%) | 0/500 (0%) | 0/500 (0%) | 0/500 (0%) | 0/500 (0%) | 0/500 (0%) | 0/500 (0%) | 0/500 (0%) | 0/500 (0%) | 0/500 (0%) | 0/500 (0%) | 3/500 (0.6%) |
| Bleeding | 3/497 (0.6%) | 1/497 (0.2%) | 0/497 (0%) | 1/497 (0.2%) | 1/497 (0.2%) | 0/497 (0%) | 0/497 (0%) | 0/497 (0%) | 0/497 (0%) | 0/497 (0%) | 0/497 (0%) | 0/497 (0%) | 1/497 (0.2%) |
| Changes in menstruation | 3/501 (0.6%) | 3/501 (0.6%) | 0/501 (0%) | 0/501 (0%) | 0/501 (0%) | 0/501 (0%) | 0/501 (0%) | 0/501 (0%) | 1/501 (0.2%) | 0/501 (0%) | 0/501 (0%) | 0/501 (0%) | 2/501 (0.4%) |
| Loss of taste | 2/469(0.43%) | 2/469 (0.43%) | 0/469 (0%) | 0/469 (0%) | 0/469 (0%) | 0/469 (0%) | 0/469 (0%) | 0/469 (0%) | 0/469 (0%) | 0/469 (0%) | 0/469 (0%) | 0/469 (0%) | 2/469 (0.43%) |
| Tingling feeling/ “pins and needles“ | 2/472 (0.42%) | 2/472 (0.42%) | 0/472 (0%) | 1/472 (0.21%) | 0/472 (0%) | 0/472 (0%) | 0/472 (0%) | 0/472 (0%) | 1/472 (0.21%) | 0/472 (0%) | 0/472 (0%) | 0/472 (0%) | 0/472 (0%) |
| Weight loss | 2/500 (0.4%) | 0/500 (0%) | 0/500 (0%) | 0/500 (0%) | 0/500 (0%) | 0/500 (0%) | 1/500 (0.2%) | 0/500 (0%) | 0/500 (0%) | 0/500 (0%) | 0/500 (0%) | 0/500 (0%) | 0/500 (0%) |
| Problems swallowing or chewing | 2/499 (0.4%) | 1/499 (0.2%) | 1/499 (0.2%) | 0/499 (0%) | 0/499 (0%) | 0/499 (0%) | 0/499 (0%) | 0/499 (0%) | 0/499 (0%) | 0/499 (0%) | 0/499 (0%) | 0/499 (0%) | 1/499 (0.2%) |
| Bilateral conjunctivitis | 2/496 (0.4%) | 2/496 (0.4%) | 0/496 (0%) | 0/496 (0%) | 0/496 (0%) | 0/496 (0%) | 0/496 (0%) | 0/496 (0%) | 0/496 (0%) | 0/496 (0%) | 0/496 (0%) | 0/496 (0%) | 2/496 (0.4%) |
| Seizures/fits | 1/498 (0.2%) | 0/498 (0%) | 0/498 (NaN%) | 0/498 (NaN%) | 0/498 (NaN%) | 0/498 (NaN%) | 0/498 (NaN%) | 0/498 (NaN%) | 0/498 (NaN%) | 0/498 (NaN%) | 0/498 (NaN%) | 0/498 (NaN%) | 0/498 (NaN%) |
| Lumps or rashes (purple/pink) on toes | 1/495 (0.2%) | 1/495 (0.2%) | 0/495 (0%) | 0/495 (0%) | 0/495 (0%) | 0/495 (0%) | 0/495 (0%) | 0/495 (0%) | 0/495 (0%) | 0/495 (0%) | 0/495 (0%) | 0/495 (0%) | 1/495 (0.2%) |
| Problems speaking or communicating | 1/489 (0.2%) | 1/489 (0.2%) | 0/489 (0%) | 0/489 (0%) | 0/489 (0%) | 0/489 (0%) | 0/489 (0%) | 0/489 (0%) | 0/489 (0%) | 0/489 (0%) | 0/489 (0%) | 0/489 (0%) | 1/489 (0.2%) |
| Fainting/ blackouts | 0/497 (0%) | 0/497 (0%) | 0/497 (NaN%) | 0/497 (NaN%) | 0/497 (NaN%) | 0/497 (NaN%) | 0/497 (NaN%) | 0/497 (NaN%) | 0/497 (NaN%) | 0/497 (NaN%) | 0/497 (NaN%) | 0/497 (NaN%) | 0/497 (NaN%) |

The differing denominators used indicate missing data.

### **Table S4**. Parental perception of mood and behaviour changes in their children.

| Characteristic | Likert scale response | | | | | | | Reasons of changes | | | |
| --- | --- | --- | --- | --- | --- | --- | --- | --- | --- | --- | --- |
|  | **1 (less)** | **2** | **3 (the same)** | **4** | **5 (more)** | **Not known** | **Other** | **Illness itself** | **Covid-19**  **pandemic** | **Both** | **Unsure** |
| Eating | 14 (2.7%) | 23 (4.5%) | 445 (86.4%) | 9 (1.7%) | 10 (1.9%) | 3 (0.6%) | 11 (2.1%) | 28 (49.1%) | 4 (7%) | 2 (3.5%) | 23 (40.4%) |
| Sleeping | 15 (2.9%) | 23 (4.5%) | 447 (86.8%) | 5 (1%) | 11 (2.1%) | 4 (0.8%) | 10 (1.9%) | 28 (52.8%) | 7 (13.2%) | 4 (7.5%) | 14 (26.4%) |
| Physical activity | 27 (5.2%) | 33 (6.4%) | 429 (83.3%) | 9 (1.7%) | 4 (0.8%) | 3 (0.6%) | 10 (1.9%) | 26 (37.7%) | 22 (31.9%) | 5 (7.2%) | 16 (23.2%) |
| Fatigue | 3 (0.6%) | 11 (2.1%) | 400 (77.8%) | 39 (7.6%) | 48 (9.3%) | 4 (0.8%) | 9 (1.8%) | 53 (53%) | 11 (11%) | 16 (16%) | 20 (20%) |
| Spending time with friends in-person | 31 (6.2%) | 27 (5.4%) | 392 (78.1%) | 19 (3.8%) | 6 (1.2%) | 17 (3.4%) | 10 (2%) | 4 (4.8%) | 66 (78.6%) | 7 (8.3%) | 7 (8.3%) |
| Spending time with friends remotely | 1 (0.2%) | 5 (1%) | 397 (80.4%) | 27 (5.5%) | 37 (7.5%) | 24 (4.9%) | 3 (0.6%) | 2 (2.8%) | 58 (81.7%) | 7 (9.9%) | 4 (5.6%) |
| Spending time watching TV, playing video/computer games, or using social media for educational purposes, including school/nursery work | 2 (0.4%) | 2 (0.4%) | 360 (71.9%) | 42 (8.4%) | 68 (13.6%) | 23 (4.6%) | 4 (0.8%) | 2 (1.8%) | 105 (92.9%) | 2 (1.8%) | 4 (3.5%) |
| Spending time watching TV, playing video/computer games, or using social media for non-educational purposes | 4 (0.8%) | 9 (1.8%) | 408 (81.8%) | 20 (4%) | 28 (5.6%) | 24 (4.8%) | 6 (1.2%) | 2 (3.4%) | 44 (75.9%) | 2 (3.4%) | 10 (17.2%) |
| Spending time outside | 36 (7.1%) | 39 (7.7%) | 364 (71.5%) | 35 (6.9%) | 18 (3.5%) | 6 (1.2%) | 11 (2.2%) | 5 (4.1%) | 89 (73%) | 11 (9%) | 17 (13.9%) |
| Attending school/nursery | 29 (5.7%) | 7 (1.4%) | 313 (61.9%) | 4 (0.8%) | 36 (7.1%) | 102 (20.2%) | 15 (3%) | 3 (3.7%) | 65 (79.3%) | 2 (2.4%) | 12 (14.6%) |
| Connectedness | 4 (0.8%) | 20 (4%) | 456 (91%) | 4 (0.8%) | 1 (0.2%) | 13 (2.6%) | 3 (0.6%) | 3 (10.7%) | 14 (50%) | 5 (17.9%) | 6 (21.4%) |
| Emotions | 11 (2.2%) | 57 (11.2%) | 411 (80.4%) | 11 (2.2%) | 3 (0.6%) | 5 (1%) | 13 (2.5%) | 24 (29.6%) | 16 (19.8%) | 10 (12.3%) | 31 (38.3%) |
| Behaviour | 5 (1%) | 37 (7.2%) | 438 (85.5%) | 8 (1.6%) | 3 (0.6%) | 5 (1%) | 16 (3.1%) | 16 (28.1%) | 7 (12.3%) | 6 (10.5%) | 28 (49.1%) |
| Relationships | 1 (0.2%) | 14 (2.8%) | 481 (95.2%) | 1 (0.2%) | 0 (0%) | 5 (1%) | 3 (0.6%) | 7 (46.7%) | 2 (13.3%) | 3 (20%) | 3 (20%) |

The differing denominators used indicate missing data.

### **Table S5**. Parental-reported mood and behaviour changes due to Covid-19 and pandemic in their children, stratified by the the effect.

| Characteristic | Caused by illness itself | | Caused by Covid-19 pandemic | |
| --- | --- | --- | --- | --- |
|  | ***Less*** | ***More*** | ***Less*** | ***More*** |
| Eating | 23/512  (4.5%) | 4/512  (0.8%) | 0  (0%) | 4/512  (0.8%) |
| Sleeping | 18/511  (3.5%) | 10/511  (2%) | 6/511  (1.2%) | 1/511  (0.2%) |
| Physical activity | 24/512  (4.7%) | 2/512  (0.4%) | 19/512  (3.7%) | 3/512  (0.6%) |
| Fatigue | 7/510  (1.4%) | 46/510  (9%) | 1/510  (0.2%) | 10/510  (2%) |
| Spending time with friends in-person | 2/485  (0.4%) | 2/485  (0.4%) | 47/485  (9.7%) | 19/485  (3.9%) |
| Spending time with friends remotely | 2/470  (0.4%) | 0  (0%) | 2/470  (0.4%) | 55/470  (11.7%) |
| Spending time watching TV, playing video/computer games, or using social media for educational purposes, including school/nursery work | 1/478  (0.2%) | 1/478  (0.2%) | 0  (0%) | 105/478  (22%) |
| Spending time watching TV, playing video/computer games, or using social media for non-educational purposes | 1/475  (0.2%) | 1/475  (0.2%) | 3/475  (0.6%) | 41/475  (8.6%) |
| Spending time outside | 5/503  (1%) | 0  (0%) | 49/503  (9.7%) | 39/503  (7.8%) |
| Attending school/nursery | 2/404  (0.5%) | 1/404  (0.3%) | 29/404  (7.2%) | 36/404  (8.9%) |
| Connectedness | 2/488  (0.4%) | 1/488  (0.2%) | 13/488  (2.7%) | 1/488  (0.2%) |
| Emotions | 22/511  (4.3%) | 2/511  (0.4%) | 13/511  (2.5%) | 2/511  (0.4%) |
| Behaviour | 14/506  (2.8%) | 1/506  (0.2%) | 0  (0%) | 7/506  (1.4%) |
| Relationships | 7/500  (1.4%) | 0  (0%) | 2/500  (0.4%) | 0  (0%) |

The differing denominators used indicate missing data.
